# Estimating Rates of Change to Interpret Quantitative Wastewater Surveillance of Disease Trends

**DOI:** 10.1101/2024.04.24.24306320

**Authors:** David A. Holcomb, Ariel Christensen, Kelly Hoffman, Allison Lee, A. Denene Blackwood, Thomas Clerkin, Javier Gallard-Góngora, Angela Harris, Nadine Kotlarz, Helena Mitasova, Stacie Reckling, Francis L. de los Reyes, Jill R. Stewart, Virginia T. Guidry, Rachel T. Noble, Marc L. Serre, Tanya P. Garcia, Lawrence S. Engel

## Abstract

**Background:** Wastewater monitoring data can be used to estimate disease trends to inform public health responses. One commonly estimated metric is the rate of change in pathogen quantity, which typically correlates with clinical surveillance in retrospective analyses. However, the accuracy of rate of change estimation approaches has not previously been evaluated.

**Objectives:** We assessed the performance of approaches for estimating rates of change in wastewater pathogen loads by generating synthetic wastewater time series data for which rates of change were known. Each approach was also evaluated on real-world data.

**Methods:** Smooth trends and their first derivatives were jointly sampled from Gaussian processes (GP) and independent errors were added to generate synthetic viral load measurements; the range hyperparameter and error variance were varied to produce nine simulation scenarios representing different potential disease patterns. The directions and magnitudes of the rate of change estimates from four estimation approaches (two established and two developed in this work) were compared to the GP first derivative to evaluate classification and quantitative accuracy. Each approach was also implemented for public SARS-CoV-2 wastewater monitoring data collected January 2021 – May 2023 at 25 sites in North Carolina, USA.

**Results:** All four approaches inconsistently identified the correct direction of the trend given by the sign of the GP first derivative. Across all nine simulated disease patterns, between a quarter and a half of all estimates indicated the wrong trend direction, regardless of estimation approach. The proportion of trends classified as plateaus (statistically indistinguishable from zero) for the North Carolina SARS-CoV-2 data varied considerably by estimation method but not by site.

**Discussion:** Our results suggest that wastewater measurements alone might not provide sufficient data to reliably track disease trends in real-time. Instead, wastewater viral loads could be combined with additional public health surveillance data to improve predictions of other outcomes.

## Introduction

The use of wastewater surveillance to monitor infectious disease expanded dramatically during the global coronavirus disease 2019 (COVID-19) pandemic, with thousands of monitoring sites active across dozens of countries by early 2023.^1^ Wastewater monitoring offers attractive features for augmenting surveillance of a wide range of pathogens and other population health-relevant targets, such as toxic metals and endogenous biomarkers, and may be particularly well-suited as an early warning system for outbreaks of novel pathogens and variants.^2–7^ However, assessing disease *trends* using wastewater surveillance faces an inherent challenge of interpretation: unlike traditional population metrics derived from counts of infected, symptomatic, or hospitalized individuals, the quantity of pathogen markers (e.g., gene targets) measured in wastewater cannot be used directly as a proxy for community disease burden. The loads of pathogen markers present in wastewater are broadly proportional to the number of infected individuals shedding the pathogen in their feces—demonstrated for SARS-CoV-2 by widely reported positive associations between wastewater viral loads and reported COVID-19 cases—but numerous biological, environmental, and site-specific factors can differentially impact measurements of wastewater pathogen loads at any given place and time.^8–12^

A common strategy for interpreting wastewater pathogen loads is to estimate traditional disease metrics like incidence rate or effective reproduction number.^12–17^ Such metrics are typically estimated by exploiting statistical associations between wastewater pathogen loads and reported cases, hospitalizations, etc. at a given site or by constructing mechanistic models of fecal shedding to estimate the number of community infections required to produce the pathogen loads measured in the community’s wastewater. These strategies require additional assumptions and data to implement, such as geographically and temporally aligned population surveillance data or pathogen-specific fecal shedding distributions. Such data, however, are often unavailable for novel pathogens and are subject to change unpredictably over the course of an outbreak or pandemic.^17–19^

An alternative strategy for interpreting wastewater pathogen loads is to assess trends over time by comparing the loads measured at different time points within the same location. Because assessing trends within-site helps to control for site-specific factors that influence pathogen load measurements, an increasing trend in wastewater measurements should correspond to an increase in infections in the community. Useful information about the direction and speed at which community infection trends are changing may therefore be inferred solely on the basis of wastewater measurements by estimating the slope of the wastewater trend at specific times, where the sign and magnitude of the slope provide the direction and rate of change, respectively.^20^

During the COVID-19 pandemic, the United States Centers for Disease Control and Prevention (CDC) described a simple regression-based approach for estimating the rate of change in SARS-CoV-2 wastewater trends over small subsets of wastewater viral load data.^21,22^ A refinement of this approach was suggested that uses reported daily COVID-19 case counts to impute wastewater viral loads on unmonitored days before applying linear regression to estimate rates of change.^20^ Both approaches produce estimated slopes (the rate of change) and associated standard errors that can be used for trend classification: a positive and statistically significant slope means the trend is increasing, a negative and significant slope means the trend is decreasing, and a slope that is not statistically significant (regardless of the sign) indicates the trend is not meaningfully changing and is classified as a plateau.^18^ Rate of change estimates from both approaches have been compared with population-based metrics (e.g., reported cases) but, to the best of our knowledge, the estimation performance and trend classification accuracy of either approach has not yet been evaluated.^18,20^

We developed a simulation-based approach to evaluate rate of change estimates using synthetic time series data for which the underlying smooth trends and their rates of change were known exactly. We sampled from Gaussian processes (GP) to jointly simulate smooth wastewater viral load trends and their first derivatives.^23–25^ Independent random errors were introduced to the simulated trends to generate synthetic measurements of wastewater viral loads, varying the smoothness of the trends and the magnitude of the errors to represent a range of potential infectious disease patterns. We evaluated four rate of change estimation approaches: the linear regression approach described by CDC, the multivariate imputation approach proposed by Al-Faliti et al. (2022), a modified univariate imputation approach requiring only wastewater measurements, and a continuous smoothing approach using generalized additive models (GAM) with numerical approximation to estimate the smooth trend and its first derivative.^20,21,26,27^ These candidate approaches were applied to the synthetic wastewater data and evaluated by comparing their rate of change estimates to the simulated GP derivatives. Finally, all four approaches were applied to public wastewater monitoring data from 25 North Carolina sewersheds to assess the impact of estimation method on the interpretation of trends in a real-world context.

## Methods

### Rate of Change Estimation Approaches

All analyses were performed in R version 4.2.2.^28,29^ R packages used are denoted by italics.

#### Rolling Regressions by Sampling Event

CDC National Wastewater Surveillance System (NWSS) recommends analyzing trends in measured wastewater viral loads by fitting simple linear regression models to a minimum of the three most-recent wastewater samples for a given location. These models use log-transformed viral load (log_10_ gene copies/day) as the response variable and date as the predictor variable.^20–22^ When fit to three observations of weekly wastewater samples or five observations of twice-weekly samples, the regression coefficient corresponds to the slope of the trend—the average daily change in viral load—over the preceding ∼15 days. The estimated rate of change can also be expressed as percent daily change (PDC), enabling more direct comparison with trends in other metrics.^21^ We estimated the rate of change on each day of sample collection by fitting rolling linear models to wastewater viral loads measured on the estimation day and the preceding four sampling events (five observations total).^21,30^ For event *i* of *N* sampling events, let 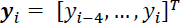 denote the log_10_-transformed wastewater viral loads and 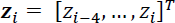 denote the sampling dates. The rate of change estimate is given by *β* in

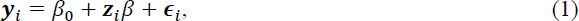

where we assume independent, normally distributed residuals 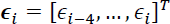.

#### Rolling Regressions on Imputed Daily Observations

##### Multivariate Imputation

While clinical and syndromic surveillance of infectious disease outcomes (e.g., incident cases, hospitalizations, and deaths) are generally reported at daily resolution, wastewater surveillance programs typically sample less frequently, often once or twice a week.^22,31^ Rate of change estimates based on small subsets of wastewater observations are subject to substantial uncertainty and temporal variability.^18,32^ To address the temporal sparsity of wastewater data, Al-Faliti et al. used daily reported cases to impute wastewater viral loads on unsampled days.^20^ Daily rates of change were estimated using rolling linear models applied over 21- or 28-day subsets of the imputed daily wastewater viral loads. Following the approach of Al-Faliti et al., we constructed five complete daily viral load datasets using the *mice* package to implement multivariate imputation using chained equations (MICE) with random forest models.^33^ We used log_10_ wastewater viral loads and a 7-day moving average of daily cases as inputs to MICE. We modified the original approach slightly by log-transforming the averaged cases for computational stability and specifying a consistent 20 iterations of the MICE algorithm for each dataset we imputed.^34^ From the five resulting complete daily datasets, we selected the realization demonstrating the highest Spearman rank correlation between 7-day average cases and the imputed daily wastewater viral loads for downstream analyses, as specified by the method developers. For the selected multivariate-imputed daily dataset, we estimated the rate of change on each original sampling day by applying the rolling linear model approach described previously to the 21 daily observations ending on the estimation day.

##### Univariate Imputation

The multivariate imputation approach relied on daily reported case data but a key motivation for estimating rates of change in wastewater surveillance data is to enable identification and interpretation of infectious disease trends using only wastewater surveillance data (i.e., when reported case data are unavailable or inadequate). We therefore also implemented a univariate time series imputation approach that used only the measured wastewater viral loads to impute viral loads on unmonitored days. Univariate imputation was conducted by Kalman smoothing on structural time series models using the *imputeTS* package.^35^ Kalman smoothing has previously been shown to flexibly estimate smooth trends in wastewater viral loads on both synthetic and various real-world wastewater surveillance data.^32^ However, as a discrete-time autoregressive approach that assumes equal-sized time steps, Kalman smoothing does not provide a direct way to estimate the rate of change and corresponding uncertainty of the modeled trend. As such, we used Kalman smoothing trend estimates to impute wastewater viral loads on unsampled days, then estimated the rate of change on each original sampling day by applying 21-day window rolling linear models to the imputed dataset.

#### First Derivatives of Smooth Functions of Time

Substantial fluctuations over short timescales in both measured wastewater viral loads and reported infections have motivated the use of a variety of smoothing approaches to better characterize infectious disease trends from noisy surveillance data.^20,26,36^ Many common smoothing techniques, including simple moving averages and locally weighted scatterplot smoothing (LOESS), use the values of neighboring observations within a user-defined window to estimate smoothed values.^17,18,20,37^ Such techniques are entirely data-dependent and do not have simple mathematical representations like those from the methods previously presented. Common time series approaches like Kalman smoothing assume equally spaced time steps and can only provide smooth estimates at discrete time points.^38,39^ By contrast, approaches that estimate continuous, smooth functions of time from the observed data can be evaluated at any arbitrary time point to obtain the corresponding estimate of the smooth trend. This feature provides a straightforward means of estimating the rate of change in the smooth trend at any moment during the monitoring period using finite differences to numerically approximate the first derivative.^38^

We used generalized additive models to estimate smoothed wastewater viral loads as continuous functions of time (see Supplemental Material, Smoothing with Generalized Additive Models). GAMs are a flexible extension of the generalized linear model that have previously been shown to provide accurate estimates of wastewater SARS-CoV-2 viral loads.^26,27,40^ We used the *mgcv* package to estimate GAMs by restricted maximum likelihood (REML) using log_10_ wastewater viral load as the response and study date as a smooth predictor term. We specified a cubic regression spline basis and the lesser of 100 or half the number of observations as the maximum basis dimension.^27,41^ Daily first derivatives and their corresponding pointwise 95% confidence intervals (CI) were estimated from GAM fits using the *gratia* package.^42^

### Simulating Differentiable Time Series

#### Simulating Smooth Trends with Known Rates of Change

We simulated wastewater trends and corresponding rates of change by sampling from Gaussian processes with squared exponential kernel covariance functions. A GP represents a distribution over all the possible smooth functions of a continuous domain (e.g., time) and is defined by its covariance function *k*(*z*_*i*_, *z*_*j*_) that relates any pair of time points *z*_*i*_, *z*_*j*_ on that domain.^23,38,43^ The time-derivative of a GP is also a GP with a covariance kernel function *k*′(*z*_*i*_, *z*_*j*_) equal to the derivative of the original covariance function with respect to times *z*_*i*_ and *z*_*j*_.^24,25^ This feature enables us to simulate both a smooth trend and its instantaneous rate of change at any finite set of time points by jointly sampling from the GP and its derivative, which follow a multivariate normal distribution (see Supplemental Material, Gaussian Process Derivatives).^25,44^

The squared exponential kernel covariance function is given by

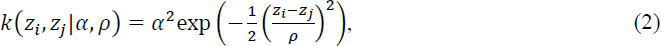

where *α* is the marginal standard deviation, a scale hyperparameter that controls the magnitude of the covariance.^25^ The rate at which correlation decays with increasing distance between times *z*_*i*_ and *z*_*j*_ is controlled by the range hyperparameter *ρ*. Smaller values of *ρ* indicate that correlation decays more quickly, so that each observation provides less information about observations at other time points. The result is a more rapidly changing, wiggly function. Conversely, larger values of *ρ* mean each observation offers greater information about its temporal neighbors, producing a more slowly changing, or smoother, trend. We implemented the squared exponential kernel and its derivatives (see Supplemental Material, Squared Exponential Kernel Function) as **R** functions and jointly sampled trend observations and derivatives using the *mvnfast* package.^44,45^

#### Generating Synthetic Wastewater Measurements

Synthetic wastewater viral load time series data (in log_10_ copies/day) were generated by jointly sampling a smooth trend and its first derivative from a GP at 1000 sequential integer locations to represent a 1000-day monitoring period with daily trend realizations. Independent random errors 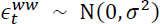 with standard deviation *σ* were independently sampled on each day *t* and added to the corresponding trend value *x_t_* to simulate the daily wastewater log_10_ viral load measurement 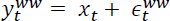. We down-sampled the synthetic wastewater measurements by selecting every third, then fourth, observation in an alternating pattern to represent a typical twice-weekly wastewater sampling frequency, denoted 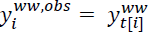 for the *i*^th^ of *N* simulated sampling events.^46^ Although our primary aim was estimating rates of change using wastewater measurements alone, implementing the multivariate imputation approach required simulating daily reported case counts that shared an underlying trend with the simulated wastewater viral loads. We sampled daily cases from a Poisson distribution with the daily log-incidence rate given by the sum of the GP trend, independent Gaussian error, and a constant mean log-incidence rate determined mechanistically for an assumed sewershed population of 200,000 (see Supplemental Material, Simulating Reported Case Counts).^12^

### Evaluating Performance of Rate of Change Estimation Approaches

#### Simulation Scenarios

We evaluated the rate of change estimation performance for three values of *ρ* so as to have varying smoothness of trends, and three values of *σ* so as to control the magnitude of variation of synthetic observations around the trend. In total, we had nine simulation scenarios. Range hyperparameter values of *ρ* = 90, *ρ* = 30, and *ρ* = 15 days were selected to produce more smooth (slowly changing), moderately smooth, and less smooth (wiggly) trends, respectively.^38^ The GP marginal standard deviation *α* = 1 was used for all simulations, such that the uncorrelated variance *σ*^2^was a quarter of the autocorrelated trend variance *α*^2^ for the less-noisy condition *σ* = 0.5; approximately half the trend variance for the moderate condition *σ* = 0.75; and equal to the trend variance for *σ* = 1. We generated 1000 synthetic datasets under each scenario and estimated the rate of change on each designated “sampling event” day (i.e., the third and seventh day of each seven-day period) by each of the four approaches (rolling linear models, multivariate imputation, univariate imputation, and generalized additive models).

Implementing an estimation approach over the entire synthetic dataset corresponds to a retrospective analysis in which previously collected data are analyzed to characterize past trends. However, active wastewater monitoring programs are primarily concerned with identifying changes in infection trends in near real-time, updating estimates as new data become available.^22^ For the rolling linear model approach these analyses are identical, as only the five most recent observations are analyzed for each estimation day. By default, the imputation and GAM approaches make use of the entire set of observations, but in real-time analyses they would be limited to only the data collected up to each estimation time point. Accordingly, we also implemented a modified local GAM approach (“rolling GAM”) that refit the GAM to only the subset of data already observed by the day for which the rate of change was being estimated. Locally restricted rolling imputations were not implemented: multiple imputation was too computationally intensive to feasibly perform across all simulation scenarios and iterations, while for univariate imputation, the maximum likelihood estimation underpinning the Kalman smoothing too often failed to converge, halting the simulations. The global imputation approaches provide upper bounds on the performance of these approaches by incorporating future information into the imputations while only using a limited window of antecedent imputed observations to estimate the rate of change on a given day.

#### Performance Metrics

For each simulated 1000-day surveillance period, we estimated the rate of change by each approach on all days with a corresponding synthetic wastewater viral load measurement after the first 90 days (a three-month baseline data collection period to accommodate the different minimum sample sizes required by each approach), for a total of 260 estimates per approach. The rate of change estimate 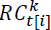 for approach *k* was compared with the sampled GP first derivative 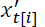 on day *t* corresponding to viral load observation *i* to assess pointwise estimation performance. The pointwise performance indicators were summarized across all estimates for a given simulated time series to calculate performance metrics for each approach. Performance metric distributions were further characterized as the median, 2.5%, and 97.5% quantiles of each metric across all 1000 simulations of each simulation scenario. Quantitative accuracy was assessed by the root mean square error, 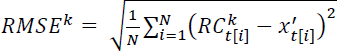.^17^ The 95% CI coverage (the proportion of 95% CIs containing the true rate of change) and average width (distance between the upper and lower 95% CI bounds) served as indicators of quantitative precision.

Each estimate was also classified as increasing or decreasing according to the sign of the point estimate (positive or negative, respectively, as the point estimate was never exactly zero).^18^ The rate of change estimate at observation *i* was considered a true positive (TP) when both the point estimate and true rate of change were positive 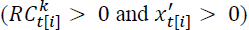 ; a true negative (TN) for 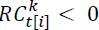 and 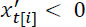 ; a false positive (FP) for 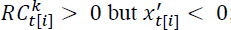; and a false negative (FN) for 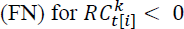 but 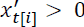. We assessed binary classification performance by sensitivity, the proportion of true increasing trends correctly classified as increasing, and specificity, the proportion of true decreasing trends correctly classified as decreasing:

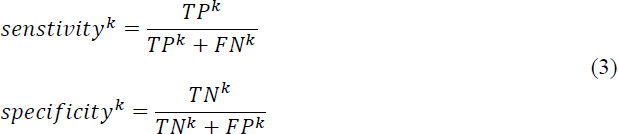

CDC suggests a third class, plateau, corresponding to low rates of change that may not warrant a response.^18,21^ In practice, however, plateaus are classified on the basis of a statistical test and identify trends with rates of change that the estimator cannot confidently differentiate from zero. Because the simulated true trend and its rate of change are known exactly, there is no directly equivalent definition available to classify true plateaus for evaluating multiclass performance. We instead incorporated the concept of varying confidence in class predictions by considering the probability that the trend was increasing. We estimated the probability of an increasing trend by computing the proportion of a rate of change estimate’s 95% CI that was greater than zero. A CI that covered only positive rates of change was assigned a 100% probability of belonging to the increasing class, while a CI that included only negative rates of change was considered to have a 0% probability of increasing. For a CI that included zero, we computed the probability of an increasing trend by dividing the rate of change value at the CI’s upper bound by the width of the CI. We performed receiver operating characteristic (ROC) curve analysis with the *yardstick* package to incorporate trend class probability into the binary classification performance assessment.^47^ Sensitivity and specificity were calculated using each observed class probability as the threshold for classifying an increasing trend, which generated an ROC curve tracing the sensitivity-specificity trade-off across probability thresholds. We used the area under the curve (AUC) to assess classification performance when treating the rate of change estimation approach as a probabilistic classifier.^48^

### Application: North Carolina SARS-CoV-2 Wastewater Viral Loads

We obtained publicly available data on wastewater SARS-CoV-2 per-capita viral loads and COVID-19 cases for 25 North Carolina sewersheds from the NC Department of Health and Human Services (NCDHHS) COVID-19 Wastewater Monitoring Dashboard.^49^ Detailed procedures for sample collection, laboratory analysis, and data processing have been described previously.^46,50^ Ten sewersheds began reporting viral loads in January 2021, with nine sewersheds added in June 2021, five more in October–November 2021, and a single addition in March 2022. We analyzed data collected through 24 May 2023, when COVID-19 case reporting ended statewide. The publicly available data listed a count of 2 for any day with 1 – 4 cases to protect privacy and no value (missing) for days with no new cases, with COVID-19 incidence reported as daily new cases per 10,000 sewershed population. We scaled by the reported sewershed population and rounded to the nearest integer to recover daily case counts, substituting 0 for missing counts and a random integer from 1 – 4 with equal probability for any recovered counts of 2. Wastewater per-capita viral loads were log_10_-transformed for all analyses, yielding units of log_10_ copies/person/day; data were provided with imputed values already substituted for non-detects, as described previously.^46,50^

For each sewershed, we estimated the rate of change on the date of each wastewater sample (after an initial 90-day baseline monitoring period) via the four estimation approaches and classified each estimate as increasing, decreasing, or plateau.^21^ We compared rate of change estimates to the first derivative of the global trend estimated by the GAM fit to the full dataset for each sewershed (“global GAM”). Agreement between the global GAM estimates and the four local estimation approaches informed only by antecedent observations was assessed using the same metrics as for the simulation study.

### Approval and Availability Statement

No human participants were involved in this research. All analyses were performed on synthetic or publicly available, aggregated data and did not require ethical approval. The code and data to perform these analyses are freely available in a permanent online repository at 10.17605/OSF.IO/BPGN4 (see Supplemental Material, Analysis Code). The original NC sewershed monitoring data may be accessed at https://covid19.ncdhhs.gov/dashboard/data-behind-dashboards.

## Results

### Simulation Study

#### Scenarios

Figure 1 presents examples of the synthetic wastewater viral load data we generated for each simulation scenario. As expected, the *ρ* = 90 days, *σ* = 0.5 log_10_ copies/day scenario produced the smoothest trend and least noisy observations, whereas the *ρ* = 15 days, *σ* = 1 log_10_ copies/day scenario produced the most wiggly trend with the noisiest observations. The trend first derivatives, corresponding to the rate of change, were influenced only by the value of *ρ* (Figure 1b). The smoothest scenarios (*ρ* = 90) produced rates of change with the smallest magnitudes, while the wigglier *ρ* = 15 scenarios produced much larger magnitude rates of change.

**Figure 1.**
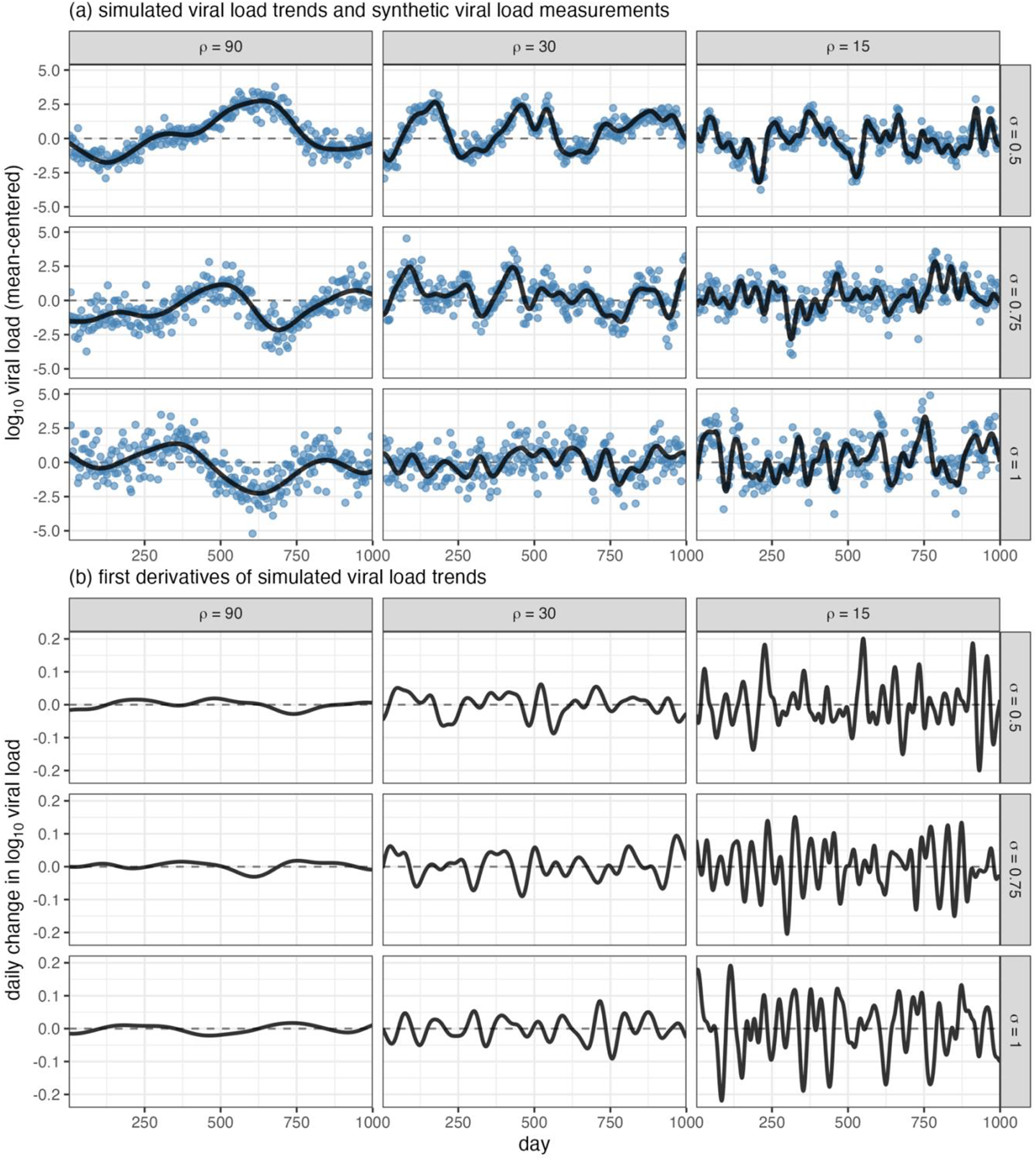
Illustrative realizations of (a) simulated wastewater viral load trend (black line) and synthetic observations (blue points) and (b) first derivative of simulated wastewater viral load trend for nine scenarios with varying specifications of the Gaussian process (GP) range parameter ρ and independent random error standard deviation *σ*

#### Rate of Change Estimates

Under the moderate *ρ* = 30 days, *σ* = 0.75 log_10_ copies/day simulation scenario, the global GAM approach generally produced smooth estimates that largely tracked both the simulated trend (Figure 2a) and its rate of change (Figure 2b). By contrast, the four local estimation approaches yielded more disjointed estimates that broadly oscillated around the true rate of change, with the rolling GAM and univariate imputation point estimates appearing to track the truth more closely and the rolling linear model swinging more dramatically between estimates. The rolling linear model estimates also exhibited the highest uncertainty, with the widest 95% CIs on average (Table S1); univariate imputation estimates typically had the narrowest CIs, which frequently did not include the true rate of change given by the GP derivative. Both the global and rolling GAM estimates generally covered the true rate of change with their 95% CIs. The rolling GAM estimates had greater uncertainty. This uncertainty resulted because each estimate was made at the extreme of the range of the observed data without the benefit of future observations to the right of the estimation point that were available to the global GAM (except for the final observation, for which the approaches, as expected, converged to identical estimates).^40^ Both imputation approaches and the rolling GAM also appeared to lag somewhat during periods of more rapid change in the trend, observable in Figure 2b as the right-ward shift in the estimated rate of change relative to the GP derivative.

**Figure 2.**
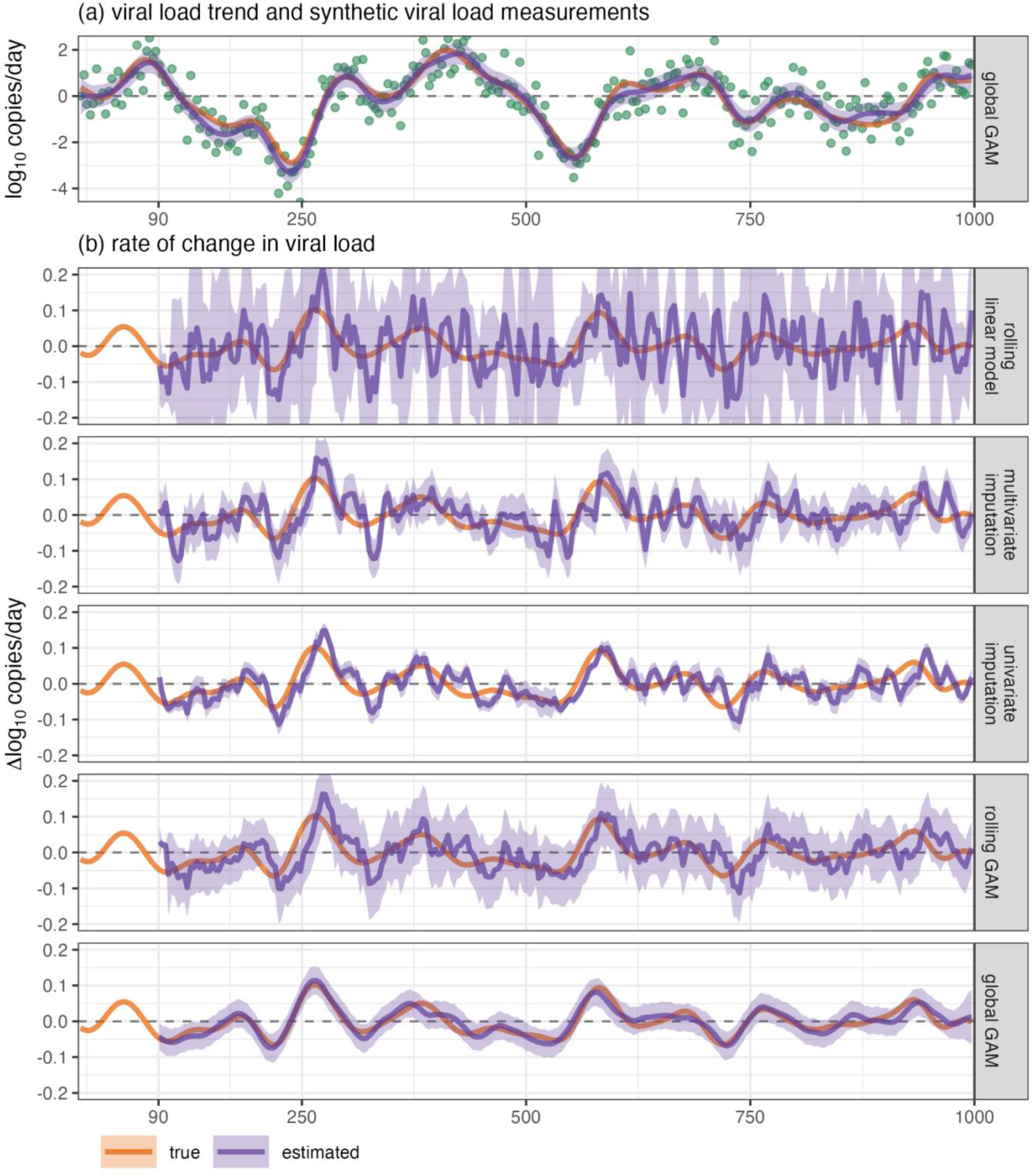
True (simulated) and estimated (a) viral load trend and (b) rate of change in viral load by each of the candidate estimation approaches for one realization of the moderately smooth, moderately noisy (*ρ* = 30, *σ* = 0.75) simulation scenario. The synthetic measurements used to fit all models are displayed as green points.

#### Estimation Performance

Across all nine scenarios, the global GAM—which utilized more data than an ongoing, real-time wastewater monitoring program would have access to—consistently produced the most accurate estimates, as indicated by lowest RMSE (Figure 3a). For the smoothest scenarios (*ρ* = 90 days), the rolling GAM exhibited similarly high accuracy regardless of the magnitude of the noise parameter *σ*, followed by the univariate imputation approach, multivariate imputation, and finally by the rolling linear model approach, which was considerably less accurate and more impacted by increasing noise variance. However, the differences in accuracy between approaches diminished for less-smooth trends as RMSE increased, such that the RMSE distributions were similar across all four local estimation approaches (median RMSE: 0.06 – 0.08 **Δ**log_10_ copies/day) for the least-smooth (*ρ* = 15) scenarios with low (*σ* = 0.5) and moderate (*σ* = 0.75) noise variance.

**Figure 3.**
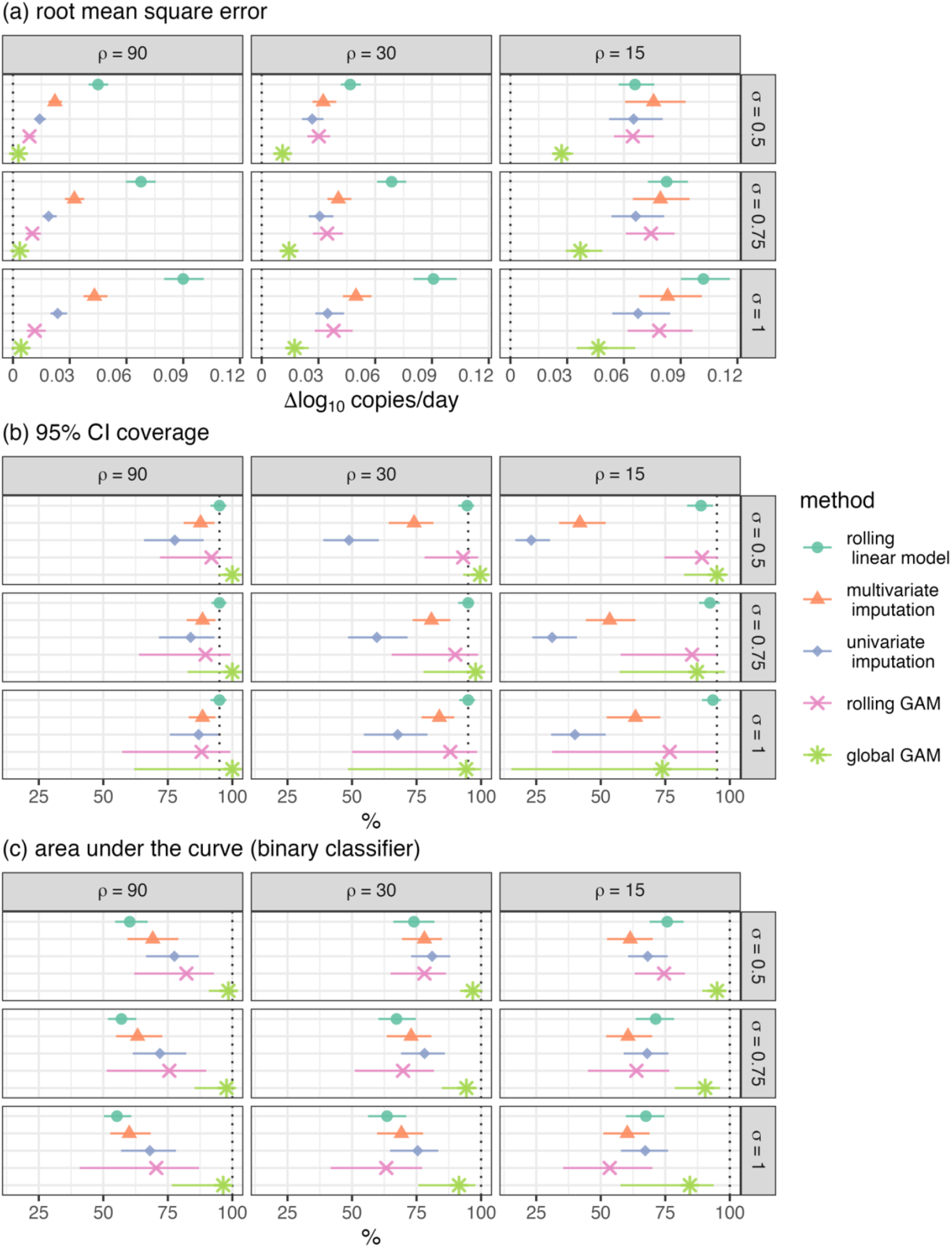
Median and 2.5^th^ – 97.5^th^ percentiles of performance metrics for candidate rate of change estimation approaches across 1000 realizations of each of the nine simulation scenarios. Vertical dotted lines indicate the target performance for each metric.

Although the rolling linear model estimates generally had the widest 95% CIs (Figure 2b), they also most consistently included the true rate of change in about 95% of intervals, the target coverage proportion (Figure 3b). The uncertainty of global GAM estimates, while much narrower than for the rolling linear model, was overly conservative, with median coverage >95% for the smoother and less noisy scenarios. Univariate imputation had coverage proportions appreciably <95% for all but the smoothest scenarios. The multivariate imputation approach also generally had coverage proportions that were <95%. The median 95% CI coverage of rolling and global GAM estimates remained relatively high across all scenarios, but both approaches demonstrated large variability in interval coverage across iterations of the more challenging (less smooth, noisier) scenarios. GAMs appear to be susceptible to estimating inappropriately smooth trends under such conditions, as observed in the essentially flat trend with narrow 95% CIs estimated by the global GAM for the *ρ* = 30, *σ* = 1 scenario in Figure S4.^26^ Such over-smoothing appears to occur more frequently when GAMs are fit to high-variance (relative to *α*) synthetic measurements generated from wiggly trends, which can produce an essentially uniform cloud of observations that envelop and obscure the trend (*ρ* = 30, *σ* = 1 scenario in Figure 1a).

The relative performance of each approach at correctly classifying the trend as increasing or decreasing was consistent across the nine simulation scenarios (Figure 3c). The global GAM consistently demonstrated the highest median AUC, although AUCs were more variable across iterations of the wigglier and noisier scenarios. Under the smoothest trend scenarios (*ρ* = 90), the rolling GAM estimates had the highest median AUC among the local estimation approaches, but also greater variability in AUC. Rolling linear models exhibited the lowest median AUC at just over 50%— only slightly better than chance. Intriguingly, the AUC of rolling linear models improved as trend smoothness decreased and the AUC of all other approaches degraded; for the wiggliest scenarios (*ρ* = 15), the rolling linear model median AUC was equal to or greater than any of the other local models. The rolling GAM approach provided the least accurate trend classifications in the least smooth, most noisy scenario, likely related to the propensity for GAMs to over-smooth under such conditions.^26^ By contrast, the univariate imputation approach performed fairly consistently across less-smooth scenarios, regardless of the noise variance, potentially due to the overly precise estimates providing more weight when the sign was correct (as was the case for the majority of estimates). However, no local estimation approach consistently achieved median sensitivities, specificities, or AUCs above 80% for any scenario (Table S1), indicating that none of these approaches would reliably classify pandemic trends as increasing or decreasing in near real-time as part of an ongoing wastewater monitoring program.

### SARS-CoV-2 Trends in North Carolina Sewersheds

The 25 NC sewersheds that we analyzed served populations ranging from 3500 – 550,000 people and were monitored over periods of 431 – 871 days, with SARS-CoV-2 viral loads reported for 121 – 245 wastewater samples at each site (Table S2). The lowest median per-capita viral loads were observed in the Wilmington sewershed, at 3.7 million copies/person/day (interquartile range [IQR] 7.6 million copies/person/day), with the highest viral loads observed at the Cary 3 site (median (IQR): 36.8 (44.3) million copies/person/day). The measured wastewater viral loads and global GAM-estimated trend, along with the corresponding rate of change estimates by each approach assessed in the simulation study, are presented separately for each sewershed in Figures S7 – S31. Concordance between the global GAM rate of change estimates and the local model estimates were similar between approaches except for rolling linear model estimates, which exhibited somewhat higher and more variable RMSE across the 25 sewersheds (Figure S6). As in the simulation study, the rolling linear model estimates had the widest 95% CIs but also included the mean global GAM estimate at approximately the target proportion of 95%. Likewise, the univariate imputation approach had the narrowest average 95% CIs, which failed to cover the majority of GAM estimates.

Under the three-class system, in which rate of change estimates are classified as increasing or decreasing only when their 95% confidence intervals exclude zero, the univariate imputation approach also consistently identified the highest proportion of estimates as clearly increasing or decreasing and the fewest as plateaus across all sites (Figure 4). The majority of estimates were classified as plateaus by each of the other approaches, while only one site (Roanoke Rapids) was majority plateau (53%) by the univariate imputation approach (Table S3). The proportion of estimates deemed plateaus was highest (often >90%) by either the rolling linear model or the rolling GAM approaches, except in Jacksonville, where the global GAM classified all but one estimate as plateau (Figure S20). Approximately three-quarters of global GAM estimates and two-thirds of multivariate imputation estimates were identified as plateaus. The relative frequency at which each estimation approach classified trends as plateaus matched between the NC data and the simulated data under the moderate (*ρ* = 30) and least smooth (*ρ* = 15) scenarios, although the global GAM produced fewer plateau calls than did the univariate imputation for the smoothest (*ρ* = 90) simulation scenarios (Figure S5). All approaches identified both increasing and decreasing trends at each site, but we observed appreciable variation between approaches in the ratio of increasing to decreasing trend classifications.

**Figure 4.**
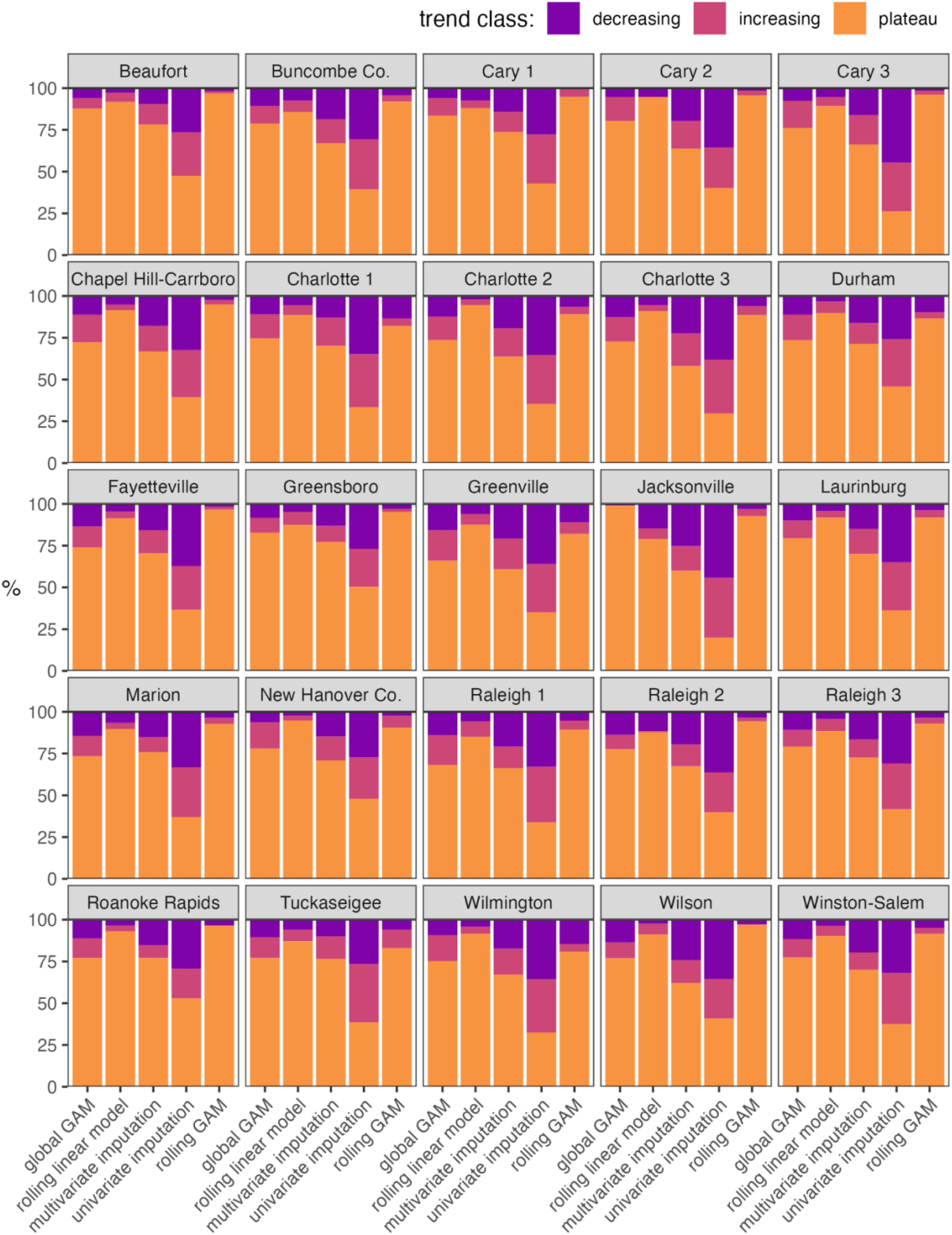
Percentage of estimates classified as decreasing (purple), increasing (mauve), or plateau (orange) trends by each rate of change estimation approach for 25 North Carolina Sewersheds.

## Discussion

Approaches for classifying viral load trends and estimating their rates of change in wastewater monitoring programs have not, to our knowledge, previously been compared for accuracy and reliability. Because trends and their slopes are not directly measured, we generated synthetic wastewater viral load time series using Gaussian processes to simulate a range of potential patterns of disease trends with known first derivatives. We implemented four rate of change estimation approaches—two previously reported and two developed herein—representing routine trend assessments from a typical, twice-weekly wastewater monitoring program that could reasonably be utilized by public health authorities without specialized statistical expertise. When applied to the synthetic time series data, all of the approaches showed only modest reliability in identifying the correct direction of the trend. The median agreement between the signs of the estimated and true rates of change typically ranged between 50% and 75%, corresponding to at least one out of every four trend assessments (i.e., once every two weeks) providing misleading conclusions about the direction of the trend. This result raises concerns about the ability to take appropriate action (e.g., issuing alerts in response to apparently increasing trends) informed by such trend classification approaches.

The addition of plateau-class trends in a three-class system partially addresses this modest reliability by requiring greater certainty before identifying trends as increasing or decreasing. As currently defined in wastewater surveillance applications, plateau status is not an inherent property of the trend but rather the product of a conventional decision criterion that the rate of change estimate was not significantly different from zero at the 5% significance level.^51^ Both the magnitude of the estimate and the precision with which it was estimated affect whether statistical significance was achieved, meaning that trends classified as plateau may have been changing too slowly to warrant attention or that the rate of change estimate was too uncertain to confidently determine the direction of change. The choice of estimation approach consistently impacted the proportion of estimates deemed plateau, though these patterns appeared mostly related to the precision of the rate of change estimates, with the inappropriately precise imputation-based approaches identifying a larger proportion of non-plateau trends. The plateau concept implies a trade-off between making a potentially incorrect determination (e.g., classifying as decreasing a trend that is truly increasing) against failing to make a determination (i.e., classifying as plateau) when conditions are in fact meaningfully changing. However, by conflating two distinct concerns—the magnitude of change and estimation uncertainty—the existing plateau definition does not directly address key questions of what is a meaningful rate of change to warrant further attention and how to balance the costs of incorrect action vs. inaction when meaningful changes in the trend are underway. Such considerations may be informally addressed by selecting more or less aggressive classification approaches or by varying the significance threshold. Greater transparency would be afforded by explicitly specifying the threshold above which rates of change would be considered meaningful, as when Keshaviah et al. specified the doubling of wastewater viral quantities as a component of an algorithm to detect infection surges.^36^ As of December 2023, CDC NWSS similarly published fixed categories defined by the percent change in viral load over the previous 15 days to classify trends, with changes ≥ 100% considered “large” increases.^52,53^

Both quantitative and classification accuracy differed between approaches, but the differences were generally observed only for smoother trends. As trends oscillated more rapidly with decreasing *ρ*, method performance degraded such that all were essentially equally poor. This dependence on characteristics of the trend itself suggests that even highly performing approaches may not be suitable in all contexts and anticipated trend smoothness should be considered when selecting a rate of change estimation approach to apply at a given monitoring site and time. While trend smoothness may be directly characterized by estimating GP range hyperparameters from previously collected monitoring data, fitting GPs is non-trivial—particularly estimating the weakly identified covariance function hyperparameters—and may be generally infeasible outside academic settings.^43,54,55^ Informal smoothness assessments may prove sufficient, for example by visually comparing global GAM-estimated trends to representative GP simulations across a range of *ρ* values. Both strategies rely on the assumption that future trends will be similar to those already observed, for which adequate data to characterize trend smoothness may be unavailable, particularly after the emergence of a novel pathogen.

To aid interpretability, we simulated wastewater trends using a simple GP with zero global mean and a squared exponential covariance function with constant marginal standard deviation *α* and a single temporal range hyperparameter *ρ* across the entire simulated monitoring period. These conditions do not fully reflect the real-world complexity of infectious disease trends. However, the flexibility of GPs readily allows extension of our simulation approach to represent smooth trends of far greater complexity. For example, the GP may be specified with multiple additive Matérn covariance functions (of which the squared exponential is a special case) to introduce fluctuations at multiple time scales, or with alternative covariance structures that represent specific physical processes (though non-Matérn functions may not be readily differentiable).^23,43,46,56,57^

The broad utility of wastewater surveillance, particularly in detecting emerging pathogen lineages, has been widely demonstrated.^58–60^ However, effectively leveraging wastewater measurements alone to estimate the rate of change of disease trends in real-time (i.e., on the day of the most recent measurement) has proved particularly challenging. While retrospective estimation of temporal trends and their rates of change by global GAMs was reasonably accurate in our simulation study and credible when applied to observed SARS-CoV-2 viral loads in NC sewersheds, real-time estimates are made at the extreme range of the data where uncertainty is greatest (as illustrated by the rolling GAM’s much noisier estimates and wider CIs relative to the global GAM). Coupling the most recent viral load measurement with recent estimated trend values provides a good indication of the neighborhood of potential values the current trend may take, but is less informative about where the trend is heading.^61^ Rather than attempting to estimate instantaneous rates of change, relative metrics with more retrospective features, such as the more recently developed CDC NWSS Wastewater Viral Activity Level metric that compares current viral loads to a long-running, site-specific baseline value, may offer a more reliable basis for understanding community disease burdens using only wastewater data.^62^ As wastewater surveillance data become increasingly relied on, any such wastewater-only metric must be thoroughly evaluated before being used to identify public health-relevant differences in community disease burden. We suggest that additional studies focus on using wastewater measurements in context with other public health metrics, such as hospitalizations or emergency department visits, to enhance predictions and updating current models to adapt to changing disease dynamics.^63^

## Data Availability

The code and data to perform these analyses are freely available in a permanent online repository at https://doi.org/10.17605/OSF.IO/BPGN4. The original NC sewershed monitoring data may be accessed at https://covid19.ncdhhs.gov/dashboard/data-behind-dashboards.

https://doi.org/10.17605/OSF.IO/BPGN4

## Acknowledgment

We gratefully acknowledge the assistance of Mitham Al-Faliti and Jeseth Delgado Vega in implementing the multivariate imputation approach. Megan Lott and Joe Brown provided insightful commentary. This work was supported by the CDC National Wastewater Surveillance System through the Epidemiology and Laboratory Capacity Cooperative Agreement with North Carolina Department of Health and Human Services, with additional support from the National Institute for Occupational Health and Safety (T42OH008673) and the NSF RAPID program (project #2029866). We also thank Steven Berkowitz and all contributors to the NC Wastewater Monitoring Network, as well as its earlier incarnation as the NC Wastewater Pathogen Research Network, which received crucial support from the NC Policy Collaborative.

## Notes

### Competing Interest Statement

The authors have declared no competing interest.

### Author Declarations

No human participants were involved in this research. All analyses were performed on synthetic or publicly available, aggregated data and did not require ethical approval. The code and data to perform these analyses are freely available in a permanent online repository at https://doi.org/10.17605/OSF.IO/BPGN4 (see Supplemental Material, Analysis Code). The original NC sewershed monitoring data may be accessed at https://covid19.ncdhhs.gov/dashboard/data-behind-dashboards.

### Summary of Updates

Typographic errors corrected in abstract and author list.

