## Supplemental Material for "Estimating Rates of Change to Interpret Quantitative Wastewater Surveillance of Disease Trends"

This supplement contains 48 pages, 3 tables, and 31 figures.

### S1 Gaussian Process Derivatives

#### *S1.1 Jointly Sampling a Gaussian Process and its Derivative*

A Gaussian process (GP) represents a distribution over all the possible smooth functions of a continuous domain (e.g., time) and is defined by its covariance function that relates any pair of locations on that domain, with any finite set of observations following a multivariate normal distribution.[1–3] Sampling a GP at a finite set of locations $\boldsymbol{z}$ provides the outputs $\boldsymbol{x}$ of a single smooth function $f(\boldsymbol{z})$ evaluated at each of those points, with the function behavior determined by the covariance kernel function $k(z_i, z_j|\boldsymbol{\theta})$ with hyperparameters $\boldsymbol{\theta}$ for each pair of locations $z_i$, $z_j$:

$$\begin{aligned} \boldsymbol{x} &= f(\boldsymbol{z}) \\ f(\boldsymbol{z}) &\sim \mathcal{GP}\left(k\left(z_i, z_j|\boldsymbol{\theta}\right)\right) \end{aligned} \tag{S1}$$

The first derivative $f'(\boldsymbol{x})$ of the smooth function is also distributed as a GP with covariance kernel function $k'\left(z_i, z_j|\boldsymbol{\theta}\right)$, the derivative of the original covariance function with respect to the locations $z_i$ and $z_j$:[4–6]

$$\begin{aligned} f'(\boldsymbol{z}) &\sim \mathcal{GP}\left(k'\left(z_i, z_j|\boldsymbol{\theta}\right)\right) \\ k'\left(z_i, z_j|\theta\right) &= \frac{\partial}{\partial z_i\,\partial z_j} k\left(z_i, z_j|\boldsymbol{\theta}\right) \end{aligned} \tag{S2}$$

This feature allows values of both the smooth function $f(\boldsymbol{z})$ and its derivative $f'(\boldsymbol{z})$ to be obtained simultaneously for all locations $\boldsymbol{z}$ by sampling jointly from the GP and its derivative, enabling us to simulate both a smooth trend and its instantaneous rate of change at any finite set of time points. Designating the original covariance kernel function $k_{00}\left(z_i, z_j\right) = k\left(z_i, z_j|\boldsymbol{\theta}\right)$ and its derivative at the same locations $k_{11}\left(z_i, z_j\right) = k'\left(z_i, z_j|\boldsymbol{\theta}\right)$, the covariance between the trend value $x_i$ at location $z_i$ and the derivative value $x'_j$ at location $z_j$ is given by

$$k_{01}\left(z_i, z_j\right) = k_{10}\left(z_j, z_i\right) = \frac{\partial}{\partial z_j} k\left(z_i, z_j|\boldsymbol{\theta}\right) \tag{S3}$$

We can then sample $\boldsymbol{x}^{all} = \begin{bmatrix} \boldsymbol{x} \\ \boldsymbol{x}' \end{bmatrix}$ at positions $\boldsymbol{z}^{all} = \begin{bmatrix} \boldsymbol{z}^{x} \\ \boldsymbol{z}^{x'} \end{bmatrix}$ by defining an indicator vector $\boldsymbol{d}^{all} = \begin{bmatrix} \boldsymbol{d}^{x} = 0 \\ \boldsymbol{d}^{x'} = 1 \end{bmatrix}$, calculating the joint covariance matrix $\Sigma^{all}$ as

$$\Sigma_{ij}^{all} = \begin{cases} k_{00}(z_i, z_j), & d_i = 0, d_j = 0 \quad \text{(both normal observations)} \\ k_{01}(z_i, z_j), & d_i = 0, d_j = 1 \quad \text{(one normal, one derivative)} \\ k_{10}(z_i, z_j), & d_i = 1, d_j = 0 \quad \text{(one derivative, one normal)} \\ k_{11}(z_i, z_j), & d_i = 1, d_j = 1 \quad \text{(both derivatives)} \end{cases} \tag{S4}$$

and sampling $\boldsymbol{x}^{all}$ from a multivariate normal distribution as $\begin{bmatrix} \boldsymbol{x} \\ \boldsymbol{x}' \end{bmatrix} \sim \text{MVN}(0, \Sigma^{all})$.[4,5]

#### *S1.2 Predicting Derivatives of a Gaussian Process*

An alternative to jointly sampling from a GP and its derivative is to sample only from the GP and predict the derivative observations $\boldsymbol{x}'$ conditional on the sampled observations $\boldsymbol{x}$ at the same locations $\boldsymbol{z}$. To predict unobserved GP values $\boldsymbol{x}_{pred} = f(\boldsymbol{z}_{pred})$ using observed GP values $\boldsymbol{x}_{obs} = f(\boldsymbol{z}_{obs})$, the conditional probability distribution is also multivariate normal:

$$x_{pred} | x_{obs} \sim \text{MVN}(\mathbf{m}, \mathbf{K}) \tag{S5}$$

The prediction mean vector $\mathbf{m}$ has an analytical solution given by

$$\mathbf{m} = \mu_{pred} + (\Sigma_{01})^T (\Sigma_{00})^{-1} (x_{obs} - \mu_{obs}) \tag{S6}$$

where $\mu_{pred}$ and $\mu_{obs}$ are the means for the prediction and observation locations, respectively (which for simplicity we usually set to zero beforehand by mean-centering), $\Sigma_{01}$ is the covariance between all observed and predicted points given by the kernel function $k(z_{obs}, z_{pred})$, and $\Sigma_{00}$ is the covariance matrix between observed locations given by the kernel function $k(z_{obs}, z_{obs})$. Likewise, the prediction covariance matrix $\mathbf{K}$ is obtained analytically as

$$\mathbf{K} = \Sigma_{11} - (\Sigma_{01})^T (\Sigma_{00})^{-1} \Sigma_{01} \tag{S7}$$

for the covariance matrix $\Sigma_{11}$ between all prediction locations produced by the kernel function $k(z_{pred}, z_{pred})$.[3,7]

Predicted derivative values $\boldsymbol{x}'$ conditional on the observations $\boldsymbol{x}$ at the same locations $\boldsymbol{z}' = \boldsymbol{z}$ can be obtained as the conditional prediction mean vector **m**:

$$
\begin{aligned}
\boldsymbol{x}' &= \mathbf{m} \\
&= \mu' + (\Sigma_{01})^T(\Sigma_{00})^{-1}(\boldsymbol{x} - \mu) \\
&= 0 + (\Sigma_{01})^T(\Sigma_{00})^{-1}(\boldsymbol{x} - 0) \\
&= (\Sigma_{01})^T(\Sigma_{00})^{-1}(\boldsymbol{x}) \\
&= \left(k_{01}(\boldsymbol{z}^x, \boldsymbol{z}^{x'})\right)^T \left(k_{00}(\boldsymbol{z}^x, \boldsymbol{z}^x)\right)^{-1}(\boldsymbol{x})
\end{aligned}
\tag{S8}
$$

where $k_{00}$ is the covariance kernel function defined previously for use with pairs of normal GP observations and $k_{01}$ is the kernel function used for one normal observation and one derivative observation. As the derivative predictions are all for the same locations as the observations, there is no prediction error and it is unnecessary to evaluate **K**. To illustrate, Figure S1 presents sampled GP observations and both the jointly-sampled derivative and the predicted derivative using a squared exponential kernel covariance function with $\alpha = 1$ and $\rho = 1$. We can visually confirm that both approaches yield equivalent values of the GP derivative and that the derivative crosses zero wherever the GP has a local minimum or maximum.

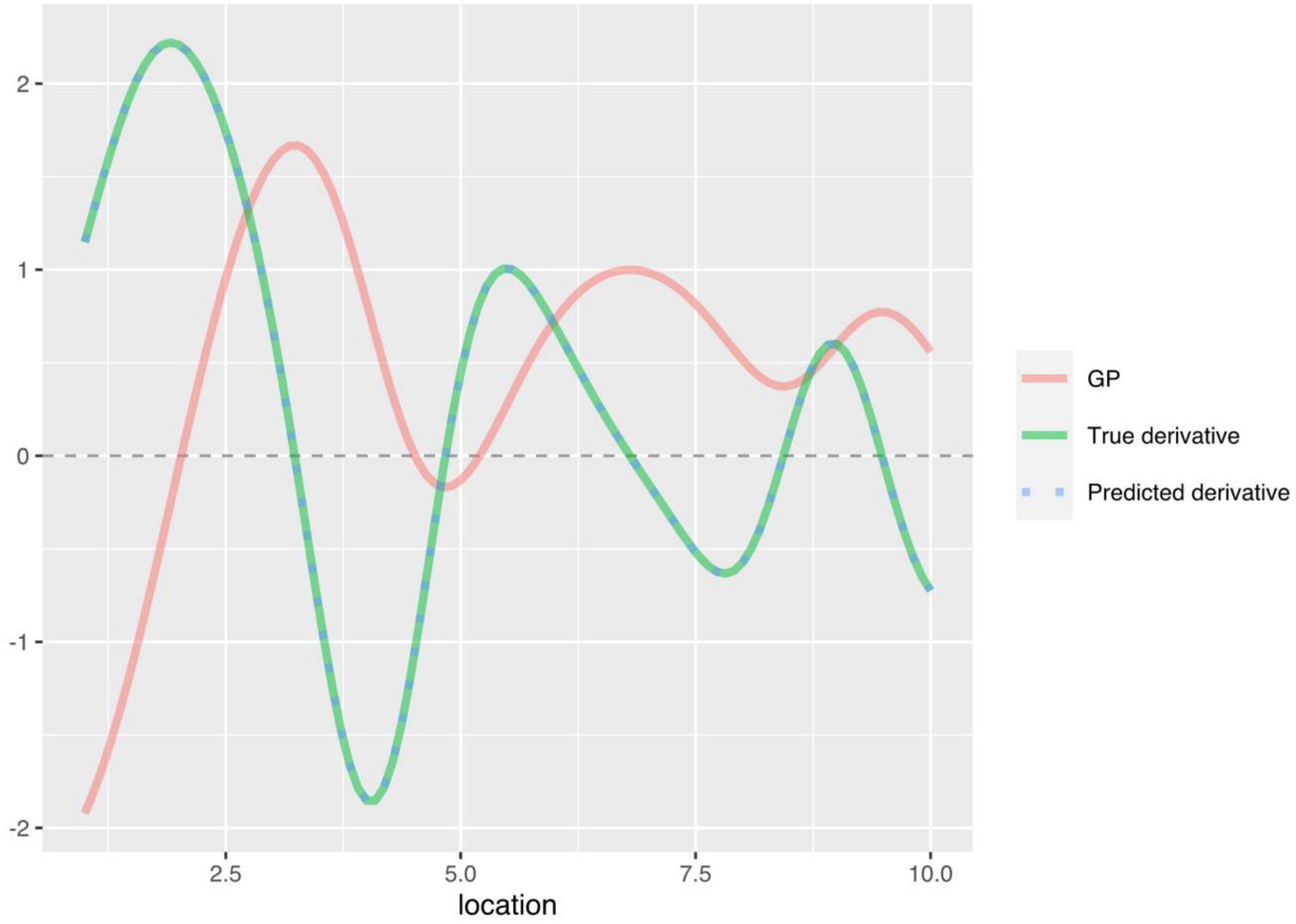

Figure S1. Comparison of jointly sampled (green solid line) and predicted (blue dotted line) derivatives of a sampled GP (red solid line)

### S2 Squared Exponential Kernel Function

The squared exponential kernel covariance function is defined as

$$k_{00}^{sqe}(z_i, z_j|\alpha, \rho) = \alpha^2 \exp\left(-\frac{1}{2}\left(\frac{z_i - z_j}{\rho}\right)^2\right) \tag{S9}$$

where $\alpha$ is the marginal standard deviation, a scale hyperparameter that controls the magnitude of the covariance.[5] For the special case $z_i = z_j$, the exponential term reduces to $\exp(0) = 1$ and $k(z_i, z_i) = \alpha^2$. The rate at which correlation decays with increasing distance between locations $z_i$ and $z_j$ is controlled by the range hyperparameter $\rho$, which corresponds to the distance between $z_i$ and $z_j$ for which the maximum covariance $\alpha^2$ has been reduced by approximately 40%. The derivative of the squared exponential kernel function $k_{11}^{sqe}(z_i, z_j|\alpha, \rho)$ for the covariance between derivative observations at locations $z_i$ and $z_j$ is given by:

$$k_{11}^{sqe} = \frac{\partial}{\partial z_i\, \partial z_j} k_{00}^{sqe}(z_i, z_j|\alpha, \rho) = \frac{\alpha^2}{\rho^4}\left(\rho^2 - (z_i - z_j)^2\right) \exp\left(-\frac{1}{2}\left(\frac{z_i - z_j}{\rho}\right)^2\right) \tag{S10}$$

and the kernel function $k_{11}^{sqe}(z_i, z_j|\alpha, \rho)$ for an observation at $z_i$ and a derivative observation at $z_j$ is:

$$k_{01}^{sqe} = \frac{\partial}{\partial z_j} k_{00}^{sqe}(z_i, z_j|\alpha, \rho) = \frac{\alpha^2}{\rho^2}(z_i - z_j) \exp\left(-\frac{1}{2}\left(\frac{z_i - z_j}{\rho}\right)^2\right) \tag{S11}$$

### S3 Smoothing with Generalized Additive Models

Smooth functions are commonly estimated using regression splines, in which polynomial regressions are fit to contiguous subsets of the observed data and constrained to have equal values, first derivatives, and second derivatives at the bounds of each subset ("knot") to produce a piecewise, smooth function over the temporal range of the data.[8–11] Introducing additional knots to subset the data more finely allows the resulting function to more closely trace the observed data, while fewer knots induce a smoother curve. Rather than manually specifying the number and locations of knots, an excess of knots may be supplied and smoothness instead induced by introducing a penalty based on the second derivative of the spline (which increases as the smoothness decreases) and an additional smoothness parameter that adjusts the strength of the penalty. Penalized splines are readily implemented as smooth predictor terms in a generalized additive model (GAM), a flexible extension of the generalized linear model that has previously demonstrated good performance estimating smooth trends in wastewater SARS-CoV-2 viral loads, among other times series applications.[8,10,12] The first derivative of each smooth predictor in a fitted GAM can be estimated for any point within the range of the data due to the continuous support of GAM-estimated smooths.[1,13] GAMs also provide a smoothness selection-corrected Bayesian covariance matrix to account for the uncertainty introduced by estimating the spline smoothness parameter, allowing confidence intervals (CIs) to be constructed for both the estimated smooth term and any estimated derivatives.[1,10]

### S4 Simulating Reported Case Counts

The multivariate imputation approach also required simulated daily case counts that shared an underlying trend with the simulated viral loads. While viral loads were simulated on a logarithmic scale with a mean of zero for simplicity (analogous to analyzing mean-centered data), case counts were simulated on the arithmetic scale by exponentiating the log scale-sampled trend, for which an additive mean on the log scale is meaningful. To simulate integer-valued case counts, we specified a sewershed population of $P = 200{,}000$ individuals and a long-run mean infection rate $\bar{\lambda} = 0.0005$ (5 cases per 10,000 population). Furthermore, we assumed only 60% of infections were reported $\left(F_{report} = 0.6\right)$ with a lag of three days after infection $(l_{report} = 3)$. These conditions yielded a mean of 100 daily new infections, which were log-transformed and added to sampled trend value $x_t$ to obtain the infection log-incidence rate $x_t^{incident}$ on study day $t$ before scaling by the reporting proportion to generate the reported log-incidence rate $x_t^{report}$. Independent Gaussian errors $\epsilon_t^{case}$ (distinct from the wastewater errors $\epsilon_t^{ww}$ but sharing the same standard deviation $\sigma$) were sampled and added to the reported log-incidence rate to obtain an over-dispersed case log-incidence rate $\ln(\lambda_t)$. Case counts were generated by sampling from a Poisson distribution with rate $\lambda_t$ and assigning the count sampled for day $t$ as the observed count three days hence to account for the specified reporting lag $l_{report}$:

$$
\begin{aligned}
y_{\mathrm{t}+l_{report}}^{case,obs} &= y_t^{case} \\
y_t^{case} &\sim \mathrm{Poisson}(\lambda_t) \\
\ln(\lambda_t) &= x_t^{report} + \epsilon_t^{case}, \qquad \epsilon_t^{case} \sim \mathrm{N}(0,\sigma^2) \\
x_t^{report} &= x_t^{incident} + \ln\left(F_{report}\right) \\
x_t^{incident} &= \mu^{incident} + x_t \\
\mu^{incident} &= \ln\left(\bar{\lambda}P\right)
\end{aligned}
\tag{S12}
$$

for the observed case count $y_{\mathrm{t}+l_{report}}^{case,obs}$ on day $t + l_{report}$. Examples of simulated cases for each of the scenarios are displayed in Figure S2, where under-reporting of cases relative to the trend is apparent.

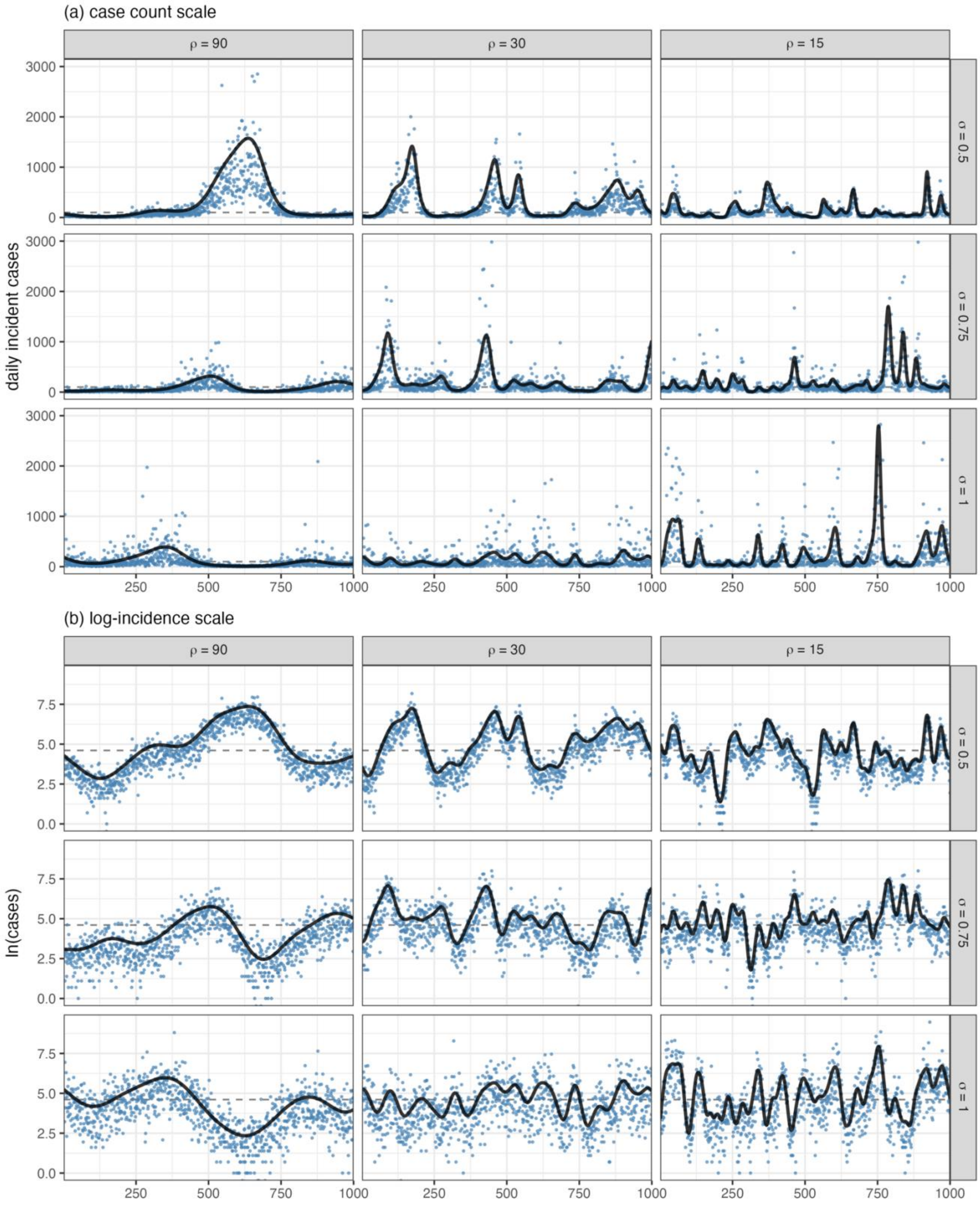

Figure S2. Synthetic reported cases simulated across nine scenarios presented on the (a) raw data scale of daily case counts and (b) natural log-incidence scale, on which the smooth trend was sampled

### S5 Indirect Simulation of Wastewater Viral Loads

When simulating wastewater measurements, the Gaussian process-sampled trend $x_t$ corresponds to log-viral load—related to the prevalence of actively shedding infections[14]—but represent log-incident infections when simulating case counts. To address this inconsistency, we also simulated wastewater viral loads using an indirect approach similar to the case simulation approach based on a consistent log-incident infections definition of the sampled GP trend. To translate incident infections into viral loads, the same daily log-incident infections $x_t^{incident}$ on day $t$ used in the case simulations were summed over an assumed 14-day average fecal shedding duration of the $D_{shed}$ days preceding each time $t$, yielding the infection log-prevalence $x_t^{prev}$. We specified that 80% of infections shed virus in feces ($F_{shed} = 0.8$) and that the average shedding infection contributed $L = 10^5$ viral gene copies/day to the wastewater viral load (that is, the virus per infection arriving at the sampling location after any dilution and loss during transport),[15] with a one-day lag $l_{shed}$ from infection onset for an indirect $\log_{10}$ wastewater viral load $y_{\mathrm{t}+l_{shed}}^{indirect,obs}$ observed on day $t = l_{shed}$:

$$
\begin{aligned}
y_{\mathrm{t}+l_{shed}}^{indirect,obs} &= y_t^{indirect} \\
y_t^{indirect} &= \log_{10}(x_t^{ww}) + \epsilon_t^{ww} \\
x_t^{ww} &= x_t^{prev} \times F_{shed} \times L \\
x_t^{prev} &= \sum_{d=0}^{D_{shed}} \exp\left(x_{\mathrm{t}-d}^{incident}\right) \\
x_t^{incident} &= \mu^{incident} + x_t \\
\mu^{incident} &= \ln\left(\bar{\lambda} P\right)
\end{aligned}
\tag{S13}
$$

Because the GP-sampled trend $x_t$ corresponds to incident infections for the indirectly simulated wastewater observations, the true trend in wastewater viral loads must also be calculated from the cumulative infections over the specified shedding duration and its first derivative is no longer known exactly. Compared with directly simulated viral load trends and observations, the scale of the indirect wastewater trends is notably compressed, relative to the variance of the synthetic observations, while maintaining the same shape (Figure S3).

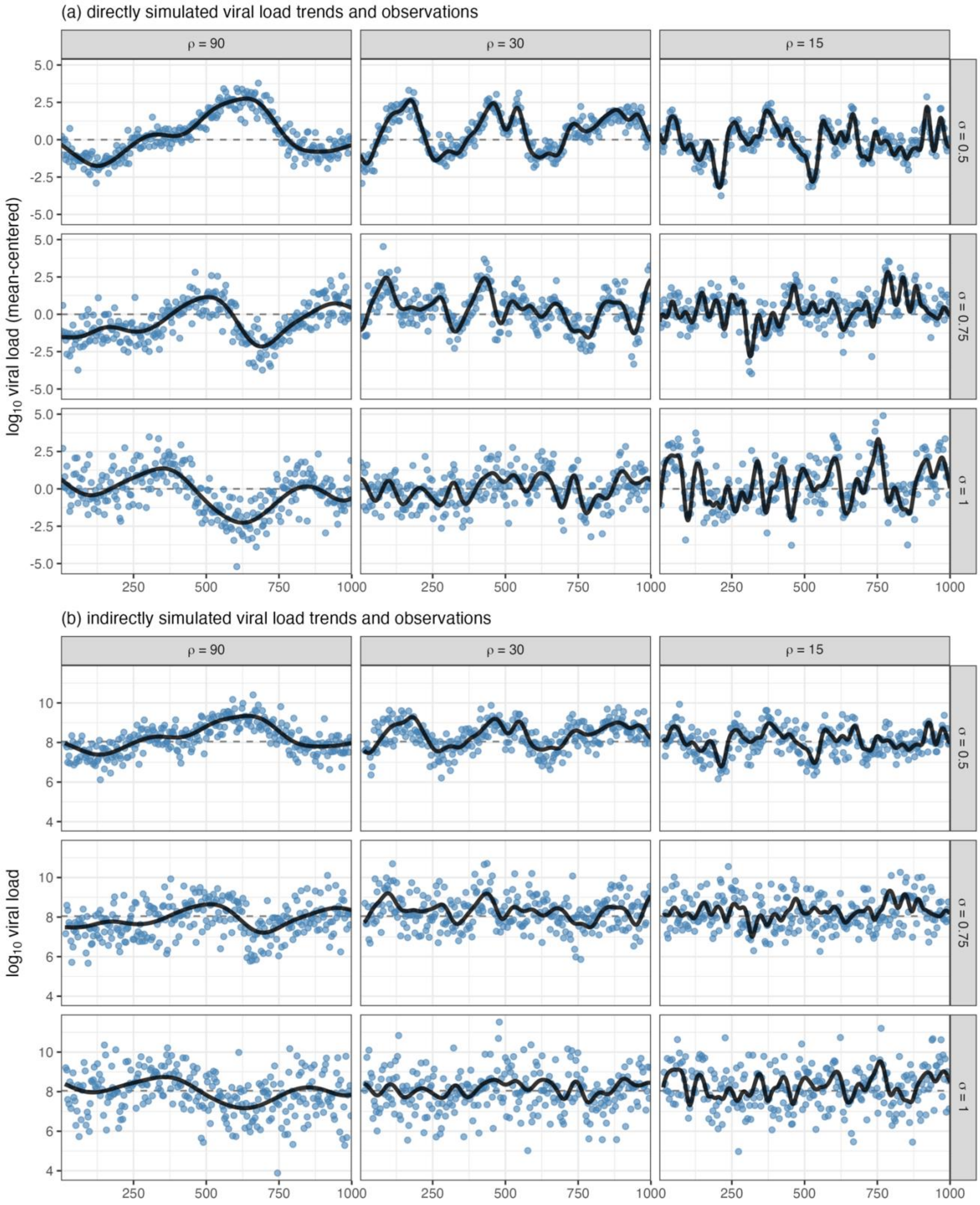

Figure S3. Comparison of wastewater trends and observations across nine simulation scenarios for (a) viral loads simulated directly from the GP-sampled trend and (b) indirectly simulated viral loads where the GP-sampled trend corresponds to log-incident infections

As our primary aim was to evaluate methods for estimating rates of change in wastewater analyte trends—independent of population infection metrics—and we simulated cases only to implement an estimation approach drawn from the literature for comparison purposes,[16] we used synthetic wastewater observations simulated directly from the sampled trend for all analyses. Indirectly simulated wastewater observations may be more appropriate for evaluating methods intended to relate wastewater measurements with population-level metrics, for example estimating the prevalence of infections or illicit drug use in a sewershed population, where the relationship between cases and wastewater is crucial and the true rate of change is of secondary interest. Such applications would likely benefit from careful consideration of the simulation parameter values assumed (e.g., global mean incidence rate, shedding proportion and duration, reporting faction and lag, etc.) to ensure realistic relationships between wastewater- and population-based synthetic data.[15]

### S6 Analysis Code

The **R** code to conduct both the simulation analysis and to produce rate of change estimates for the real-world NC wastewater SARS-CoV-2 viral load monitoring data is available at https://doi.org/10.17605/OSF.IO/BPGN4. The custom functions written to implement the analysis are provided both in an **R** script ("roc_functions.R") that can be sourced for direct use and described in detail in an HTML report ("roc_notebook.html"). Additionally, a compressed **R** project folder ("wastewaterRateOfChange.zip") is provided that contains all the data and code necessary to reproduce the HTML report. To do so, download the compressed file to a computer with recent versions (e.g., released in mid-2022 or later) of the RStudio integrated development environment (IDE) and the **R** platform installed and unzip it. Within the unzipped directory, open the "wastewaterRateOfChange.Rproj" file to launch the project in a new **R** session in RStudio. Use the "Open" dialogue within RStudio to navigate to the "roc_notebook.qmd" Quarto markdown file in the "script" subdirectory. Install any missing packages as prompted; the code can now be run interactively through the .qmd file or in its entirety using the "render" functionality in RStudio to reproduce the HTML report.

### S7 Simulation Study Rate of Change Estimates

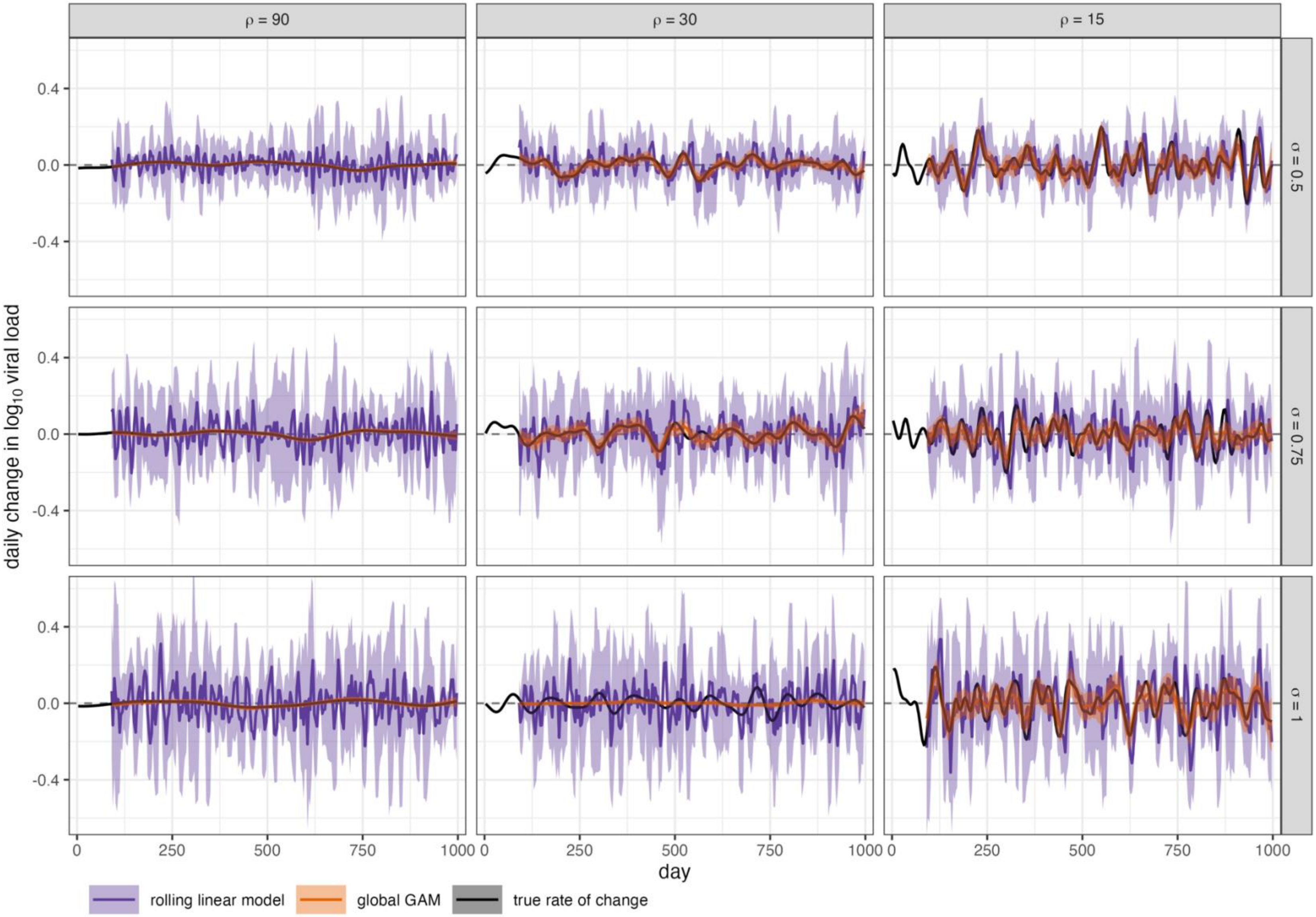

Figure S4. Mean and 95% CI estimated rates of change by the rolling linear model (purple) and GAM fit to the full dataset (orange) compared with the true first derivative of the viral load trend (black) for illustrative realizations of the nine simulation scenarios

### S8 Estimation Performance by Simulation Scenario

Table S1. Median (2.5th – 97.5th percentiles) of estimation performance across nine simulation scenarios with differing degrees of smoothness ($\rho$) and observation noise ($\sigma$)

| $\rho$ | $\sigma$ | Method | RMSE [Δlog$_{10}$ gc/day] | Mean 95% CI Width [Δlog$_{10}$ gc/day] | 95% CI Coverage [%] | Sensitivity [%] | Specificity [%] | AUC [%] |
|---|---|---|---|---|---|---|---|---|
| 90 | *Scenario 1: high smoothness, low noise* | | | | | | | |
| | 0.5 | rolling linear model | 0.04 (0.04, 0.05) | 0.27 (0.24, 0.29) | 95 (92, 98) | 58 (51, 65) | 58 (51, 65) | 60 (55, 67) |
| | | multivariate imputation | 0.02 (0.02, 0.03) | 0.07 (0.06, 0.08) | 88 (81, 93) | 64 (54, 74) | 63 (54, 74) | 69 (59, 79) |
| | | univariate imputation | 0.01 (0.01, 0.02) | 0.04 (0.03, 0.04) | 78 (66, 89) | 70 (59, 82) | 70 (59, 82) | 78 (67, 87) |
| | | rolling GAM | 0.01 (0.01, 0.01) | 0.04 (0.02, 0.05) | 92 (72, 100) | 76 (56, 89) | 75 (54, 88) | 82 (62, 93) |
| | | global GAM | 0.00 (0.00, 0.00) | 0.02 (0.01, 0.02) | 100 (95, 100) | 93 (76, 100) | 92 (75, 100) | 98 (91, 100) |
| | *Scenario 2: high smoothness, moderate noise* | | | | | | | |
| | 0.75 | rolling linear model | 0.07 (0.06, 0.08) | 0.40 (0.36, 0.44) | 95 (92, 98) | 55 (49, 62) | 55 (49, 62) | 57 (52, 63) |
| | | multivariate imputation | 0.03 (0.03, 0.04) | 0.11 (0.10, 0.12) | 88 (82, 93) | 59 (51, 69) | 59 (51, 70) | 63 (55, 73) |
| | | univariate imputation | 0.02 (0.02, 0.02) | 0.06 (0.05, 0.06) | 84 (72, 93) | 66 (55, 78) | 65 (55, 75) | 72 (61, 82) |
| | | rolling GAM | 0.01 (0.01, 0.01) | 0.04 (0.02, 0.06) | 90 (64, 99) | 71 (44, 87) | 71 (44, 87) | 76 (51, 90) |
| | | global GAM | 0.00 (0.00, 0.01) | 0.02 (0.01, 0.03) | 100 (83, 100) | 91 (69, 100) | 91 (67, 100) | 98 (85, 100) |
| | *Scenario 3: high smoothness, high noise* | | | | | | | |
| | 1 | rolling linear model | 0.09 (0.08, 0.10) | 0.53 (0.47, 0.59) | 95 (92, 98) | 54 (48, 61) | 54 (48, 60) | 55 (50, 61) |
| | | multivariate imputation | 0.04 (0.04, 0.05) | 0.14 (0.13, 0.16) | 88 (83, 93) | 57 (48, 66) | 57 (50, 65) | 60 (53, 68) |
| | | univariate imputation | 0.02 (0.02, 0.03) | 0.07 (0.07, 0.08) | 87 (76, 95) | 63 (52, 73) | 62 (53, 73) | 68 (57, 78) |
| | | rolling GAM | 0.01 (0.01, 0.02) | 0.04 (0.02, 0.07) | 88 (57, 99) | 67 (36, 86) | 67 (36, 85) | 70 (41, 87) |
| | | global GAM | 0.00 (0.00, 0.01) | 0.02 (0.01, 0.03) | 100 (62, 100) | 89 (60, 100) | 90 (60, 100) | 96 (77, 100) |
| 30 | *Scenario 4: moderate smoothness, low noise* | | | | | | | |
| | 0.5 | rolling linear model | 0.05 (0.04, 0.05) | 0.26 (0.24, 0.29) | 95 (91, 97) | 68 (60, 77) | 68 (60, 76) | 74 (66, 82) |
| | | multivariate imputation | 0.03 (0.03, 0.04) | 0.08 (0.07, 0.08) | 74 (64, 82) | 71 (62, 79) | 71 (62, 80) | 78 (69, 85) |
| | | univariate imputation | 0.03 (0.02, 0.03) | 0.04 (0.03, 0.04) | 49 (39, 60) | 75 (66, 84) | 75 (65, 84) | 81 (73, 88) |
| | | rolling GAM | 0.03 (0.02, 0.04) | 0.12 (0.08, 0.15) | 93 (78, 99) | 71 (58, 80) | 71 (58, 81) | 78 (65, 86) |
| | | global GAM | 0.01 (0.01, 0.01) | 0.06 (0.04, 0.07) | 100 (93, 100) | 90 (80, 97) | 90 (79, 97) | 97 (92, 99) |
| | *Scenario 5: moderate smoothness, moderate noise* | | | | | | | |
| | 0.75 | rolling linear model | 0.07 (0.06, 0.08) | 0.40 (0.36, 0.44) | 95 (91, 97) | 63 (55, 71) | 63 (56, 70) | 67 (60, 75) |

| *ρ* | *σ* | Method | RMSE [$\Delta\log_{10}$ gc/day] | Mean 95% CI Width [$\Delta\log_{10}$ gc/day] | 95% CI Coverage [%] | Sensitivity [%] | Specificity [%] | AUC [%] |
|---|---|---|---|---|---|---|---|---|
| | | multivariate imputation | 0.04 (0.03, 0.05) | 0.11 (0.10, 0.12) | 81 (73, 88) | 67 (57, 75) | 67 (57, 75) | 73 (63, 81) |
| | | univariate imputation | 0.03 (0.02, 0.04) | 0.05 (0.05, 0.06) | 60 (48, 72) | 72 (61, 81) | 71 (62, 80) | 78 (69, 86) |
| | | rolling GAM | 0.03 (0.03, 0.04) | 0.12 (0.07, 0.18) | 90 (65, 99) | 65 (48, 77) | 65 (47, 76) | 70 (51, 82) |
| | | global GAM | 0.01 (0.01, 0.02) | 0.06 (0.04, 0.08) | 98 (78, 100) | 86 (72, 95) | 86 (72, 95) | 94 (85, 98) |
| | *Scenario 6: moderate smoothness, high noise* | | | | | | | |
| | 1 | rolling linear model | 0.09 (0.08, 0.10) | 0.53 (0.48, 0.59) | 95 (92, 98) | 60 (53, 68) | 60 (53, 67) | 64 (56, 71) |
| | 1 | multivariate imputation | 0.05 (0.04, 0.06) | 0.15 (0.13, 0.16) | 84 (77, 90) | 64 (55, 73) | 64 (55, 72) | 69 (60, 78) |
| | 1 | univariate imputation | 0.03 (0.03, 0.04) | 0.07 (0.07, 0.08) | 68 (55, 79) | 69 (59, 78) | 69 (59, 78) | 75 (65, 83) |
| | 1 | rolling GAM | 0.04 (0.03, 0.05) | 0.13 (0.06, 0.20) | 88 (50, 98) | 60 (40, 73) | 60 (41, 72) | 63 (42, 77) |
| | 1 | global GAM | 0.02 (0.01, 0.02) | 0.07 (0.03, 0.09) | 94 (48, 100) | 83 (64, 94) | 83 (65, 94) | 91 (76, 98) |
| 15 | *Scenario 7: low smoothness, low noise* | | | | | | | |
| 15 | 0.5 | rolling linear model | 0.07 (0.06, 0.08) | 0.27 (0.24, 0.30) | 89 (83, 93) | 70 (62, 77) | 69 (62, 77) | 76 (69, 82) |
| 15 | 0.5 | multivariate imputation | 0.08 (0.06, 0.09) | 0.09 (0.08, 0.10) | 42 (34, 52) | 58 (49, 67) | 58 (49, 68) | 61 (52, 70) |
| 15 | 0.5 | univariate imputation | 0.07 (0.05, 0.08) | 0.04 (0.04, 0.04) | 23 (17, 30) | 65 (57, 73) | 65 (57, 73) | 68 (61, 76) |
| 15 | 0.5 | rolling GAM | 0.06 (0.05, 0.08) | 0.21 (0.15, 0.26) | 89 (75, 95) | 68 (57, 77) | 68 (57, 77) | 75 (63, 83) |
| 15 | 0.5 | global GAM | 0.03 (0.02, 0.03) | 0.11 (0.08, 0.12) | 95 (82, 99) | 87 (78, 94) | 87 (78, 94) | 95 (89, 98) |
| 15 | *Scenario 8: low smoothness, moderate noise* | | | | | | | |
| 15 | 0.75 | rolling linear model | 0.08 (0.07, 0.09) | 0.40 (0.36, 0.44) | 92 (88, 96) | 66 (58, 73) | 66 (58, 74) | 71 (64, 78) |
| 15 | 0.75 | multivariate imputation | 0.08 (0.06, 0.09) | 0.12 (0.11, 0.13) | 53 (44, 63) | 58 (49, 67) | 58 (50, 66) | 61 (52, 70) |
| 15 | 0.75 | univariate imputation | 0.07 (0.05, 0.08) | 0.06 (0.05, 0.06) | 31 (23, 41) | 64 (55, 73) | 64 (55, 73) | 68 (59, 76) |
| 15 | 0.75 | rolling GAM | 0.07 (0.06, 0.09) | 0.22 (0.12, 0.30) | 85 (58, 95) | 60 (44, 71) | 60 (44, 71) | 64 (45, 77) |
| 15 | 0.75 | global GAM | 0.04 (0.03, 0.05) | 0.11 (0.07, 0.14) | 87 (57, 98) | 82 (68, 91) | 82 (69, 91) | 90 (79, 96) |
| 15 | *Scenario 9: low smoothness, high noise* | | | | | | | |
| 15 | 1 | rolling linear model | 0.10 (0.09, 0.12) | 0.53 (0.48, 0.59) | 93 (89, 97) | 63 (56, 71) | 63 (56, 71) | 67 (60, 75) |
| 15 | 1 | multivariate imputation | 0.08 (0.07, 0.10) | 0.15 (0.14, 0.17) | 63 (52, 73) | 57 (48, 66) | 58 (48, 67) | 60 (51, 69) |
| 15 | 1 | univariate imputation | 0.07 (0.05, 0.08) | 0.07 (0.07, 0.08) | 40 (31, 52) | 63 (54, 72) | 63 (54, 73) | 67 (58, 76) |
| 15 | 1 | rolling GAM | 0.08 (0.06, 0.10) | 0.20 (0.07, 0.32) | 77 (31, 95) | 53 (36, 67) | 53 (36, 67) | 53 (35, 70) |
| 15 | 1 | global GAM | 0.05 (0.03, 0.07) | 0.10 (0.02, 0.15) | 74 (15, 95) | 76 (54, 88) | 76 (52, 88) | 85 (58, 94) |

### S9 Plateau Classification by Simulation Scenario

Because the simulated true trend and its rate of change are known exactly, there is no directly equivalent definition available to classify true plateaus for the simulated trend to evaluate multiclass performance. However, applying the standard plateau definition (non-significant slope at a 5% significance level) to the simulation study estimates, we observed notable differences between methods in the proportion of estimates classified as plateaus, particularly under the less-smooth scenarios (Figure S5).

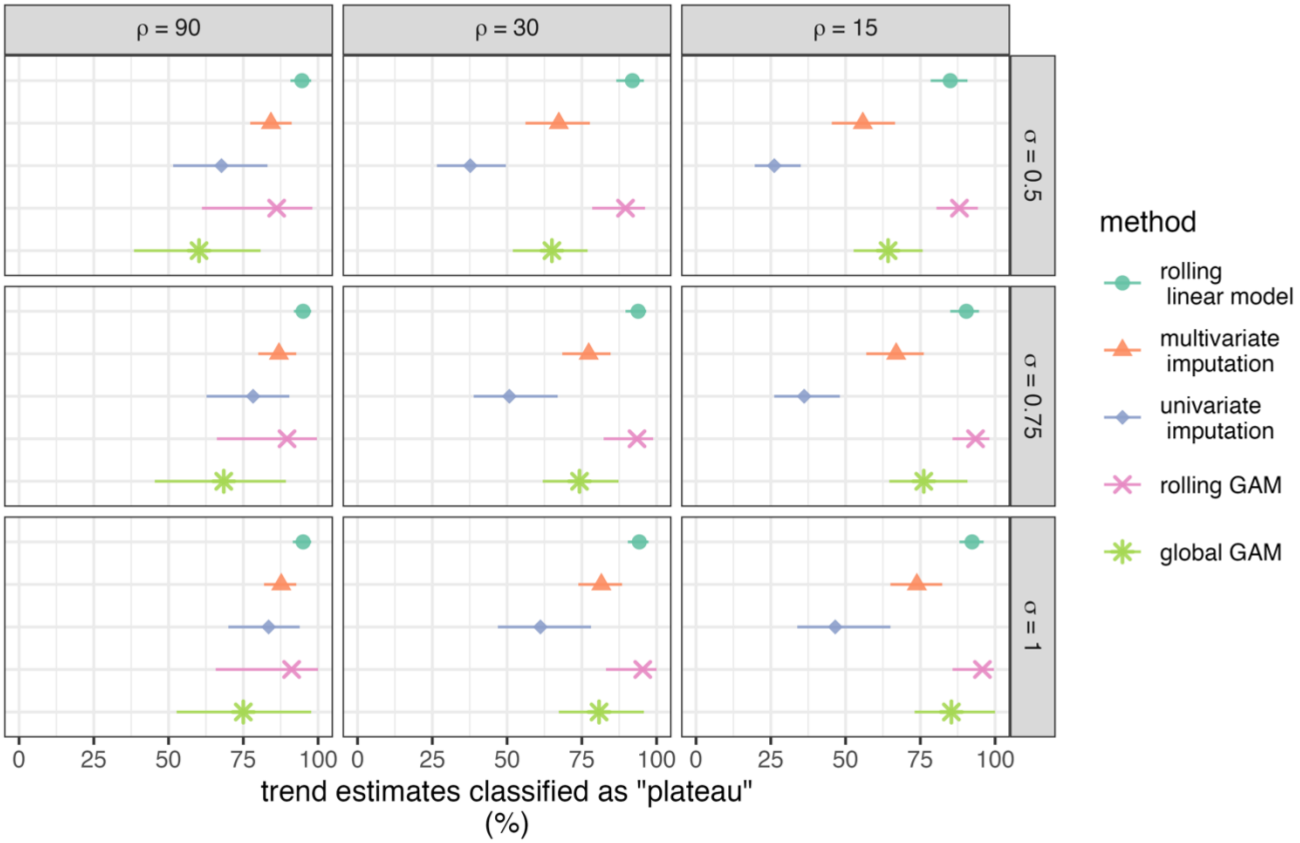

Figure S5. Median and 2.5$^{th}$ – 97.5$^{th}$ percentiles of the percentage of estimates classified as plateau (not significantly different from zero at the 5% significance level) by each rate of change estimation method across 1000 realizations of each of the nine simulation scenarios.

### S10 North Carolina Sewershed Characteristics

Table S2. Characteristics of North Carolina Sewersheds

| Sewershed | County | Population Served | Monitoring Start Date | Days Monitored[a] | Samples Reported | Median (IQR) Daily Reported Cases | Median (IQR) Per-Capita Viral Load, million gene copies/person/day |
|---|---|---|---|---|---|---|---|
| Beaufort | Carteret | 3500 | 2021-01-03 | 871 | 245 | 1 (3) | 6.7 (18.5) |
| Buncombe Co. | Buncombe, Henderson | 173000 | 2021-06-19 | 704 | 185 | 36 (48) | 9.9 (11.3) |
| Cary 1 | Wake | 84189 | 2021-11-16 | 554 | 157 | 13 (23) | 12.8 (18.5) |
| Cary 2 | Wake | 74331 | 2021-11-16 | 554 | 156 | 9 (18) | 17.5 (17.9) |
| Cary 3 | Wake | 75886 | 2021-11-16 | 554 | 149 | 15 (30) | 36.8 (44.3) |
| Chapel Hill - Carrboro | Orange | 78141 | 2021-01-06 | 868 | 238 | 11 (17) | 6.3 (11.7) |
| Charlotte 1 | Mecklenburg | 68685 | 2021-01-03 | 871 | 226 | 14 (22) | 11.3 (15.3) |
| Charlotte 2 | Mecklenburg | 182501 | 2021-01-03 | 871 | 226 | 27 (39) | 10.0 (14.6) |
| Charlotte 3 | Mecklenburg | 120000 | 2021-06-01 | 722 | 189 | 25 (38) | 17.9 (24.4) |
| Durham | Durham | 108105 | 2021-01-06 | 868 | 240 | 16 (25) | 8.6 (14.5) |
| Fayetteville | Cumberland | 151589 | 2021-06-19 | 704 | 198 | 33 (49) | 10.1 (11.0) |
| Greensboro | Guilford | 135821 | 2021-06-18 | 705 | 193 | 25 (34) | 9.8 (14.0) |
| Greenville | Pitt | 89616 | 2021-01-03 | 871 | 242 | 17 (26) | 7.1 (11.8) |
| Jacksonville | Onslow | 41819 | 2022-03-19 | 431 | 121 | 5 (9) | 5.7 (8.9) |
| Laurinburg | Scotland | 15527 | 2021-06-17 | 706 | 186 | 3 (6) | 6.9 (12.0) |
| Marion | McDowell | 8459 | 2021-06-17 | 706 | 191 | 2 (4) | 5.6 (10.7) |
| New Hanover Co. | New Hanover | 67743 | 2021-01-22 | 852 | 235 | 7 (12) | 5.0 (10.5) |
| Raleigh 1 | Wake | 550000 | 2021-01-06 | 868 | 231 | 100 (146) | 7.0 (10.6) |
| Raleigh 2 | Wake | 30655 | 2021-10-21 | 580 | 160 | 8 (15) | 10.8 (15.1) |
| Raleigh 3 | Wake | 7776 | 2021-10-21 | 580 | 158 | 2 (4) | 15.8 (25.8) |
| Roanoke Rapids | Halifax, Northampton | 14320 | 2021-06-19 | 704 | 194 | 4 (7) | 9.3 (14.2) |
| Tuckaseigee | Jackson | 13296 | 2021-01-03 | 871 | 213 | 2 (3) | 4.0 (8.1) |
| Wilmington | New Hanover | 58361 | 2021-01-05 | 869 | 235 | 9 (15) | 3.7 (7.6) |
| Wilson | Wilson | 49384 | 2021-06-19 | 704 | 194 | 8 (14) | 6.2 (12.1) |
| Winston-Salem | Forsyth | 178000 | 2021-06-19 | 704 | 186 | 38 (54) | 5.3 (6.2) |

[a] monitoring ended for all sewersheds on 2023-05-24

### S11 Relative Performance of Estimation Methods in NC Sewersheds

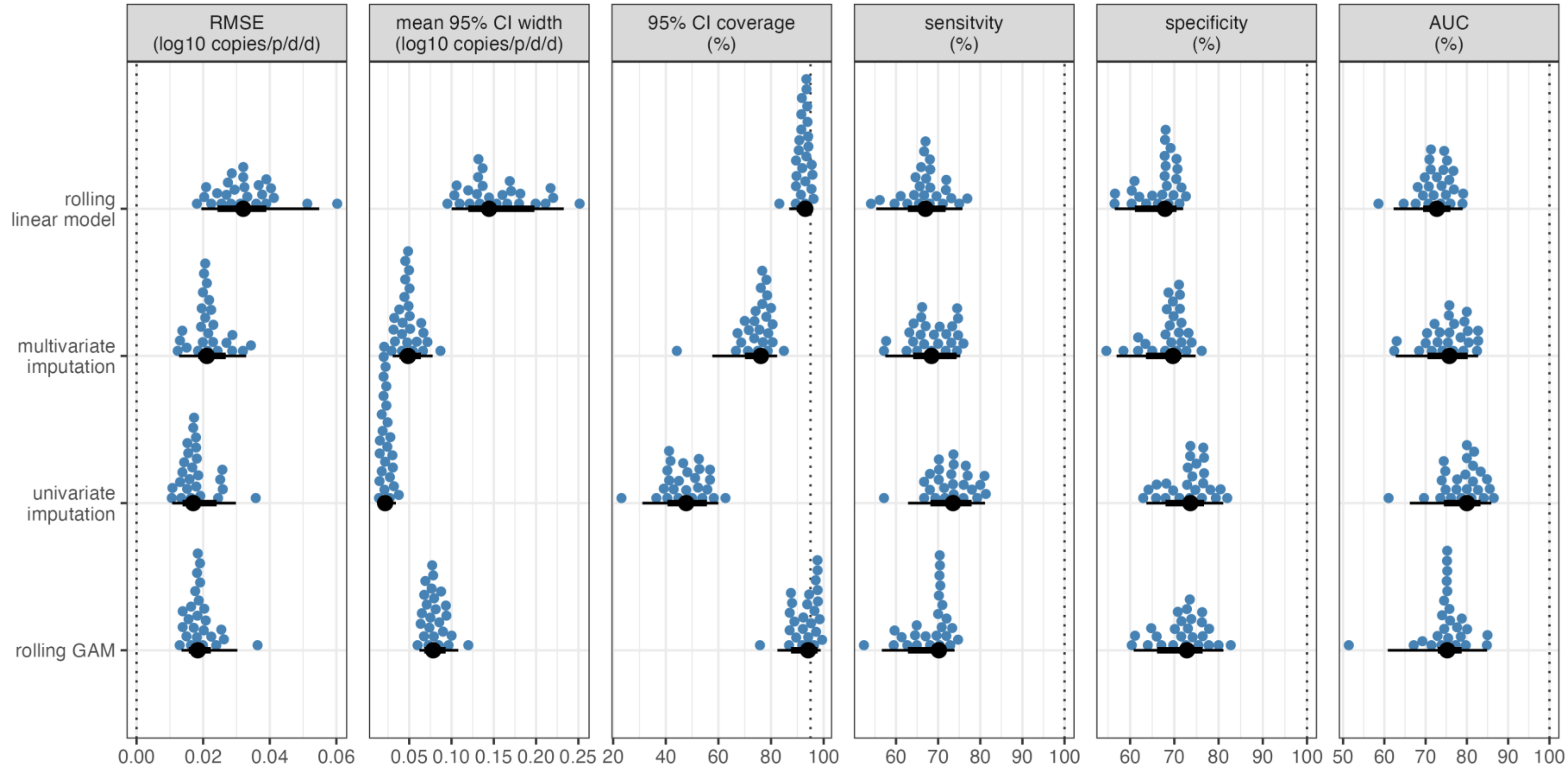

Figure S6. Comparison of rate of change estimates from local estimation methods to global GAM estimates for SARS-CoV-2 wastewater viral loads at 25 NC sewersheds. Each blue point corresponds to a sewershed, with the black point and line presenting the median and 2.5$^{th}$ – 97.5$^{th}$ percentiles of the performance metric across sewersheds for a given local estimation method. Vertical dotted lines indicate the target performance for each metric

### S12 Trend Classifications in NC Sewersheds

Table S3. Percentage of rate of change estimates classified as positive (two class) and increasing, decreasing, or plateau (three class) in North Carolina sewersheds by estimation method

| Sewershed | N | Positive Slope (% of estimates) | | | | | Slope Classification[a] (% of estimates) | | | | | | | | | | | | | | |
|---|---|---|---|---|---|---|---|---|---|---|---|---|---|---|---|---|---|---|---|---|---|
| | | | | | | | Global GAM | | | RLM | | | MI | | | UI | | | Rolling GAM | | |
| | | Global GAM | RLM | MI | UI | Rolling GAM | ↑ | ↓ | — | ↑ | ↓ | — | ↑ | ↓ | — | ↑ | ↓ | — | ↑ | ↓ | — |
| Beaufort | 219 | 51 | 49 | 52 | 52 | 49 | 6 | 6 | 88 | 5 | 3 | 92 | 12 | 10 | 78 | 26 | 26 | 47 | 1 | 2 | 97 |
| Buncombe Co. | 160 | 44 | 51 | 51 | 49 | 48 | 11 | 11 | 79 | 7 | 8 | 86 | 14 | 19 | 67 | 30 | 31 | 39 | 4 | 4 | 92 |
| Cary 1 | 133 | 43 | 50 | 44 | 49 | 43 | 11 | 6 | 83 | 5 | 8 | 88 | 12 | 14 | 74 | 29 | 28 | 43 | 5 | 1 | 95 |
| Cary 2 | 132 | 34 | 45 | 44 | 42 | 36 | 14 | 5 | 80 | 0 | 5 | 95 | 17 | 20 | 64 | 24 | 36 | 40 | 3 | 2 | 95 |
| Cary 3 | 130 | 36 | 44 | 47 | 42 | 42 | 16 | 8 | 76 | 5 | 5 | 89 | 18 | 16 | 66 | 29 | 45 | 26 | 2 | 2 | 96 |
| Chapel Hill - Carrboro | 213 | 43 | 47 | 49 | 48 | 46 | 16 | 11 | 72 | 3 | 5 | 92 | 15 | 18 | 67 | 28 | 32 | 39 | 3 | 2 | 95 |
| Charlotte 1 | 201 | 48 | 48 | 51 | 48 | 45 | 14 | 11 | 75 | 6 | 5 | 89 | 17 | 13 | 70 | 32 | 35 | 33 | 4 | 13 | 82 |
| Charlotte 2 | 201 | 45 | 49 | 51 | 48 | 44 | 14 | 12 | 74 | 3 | 2 | 95 | 17 | 19 | 64 | 29 | 35 | 35 | 4 | 6 | 89 |
| Charlotte 3 | 165 | 49 | 46 | 47 | 48 | 48 | 15 | 13 | 73 | 4 | 5 | 91 | 19 | 22 | 58 | 32 | 38 | 30 | 5 | 6 | 88 |
| Durham | 216 | 46 | 52 | 48 | 50 | 46 | 15 | 11 | 74 | 7 | 3 | 90 | 12 | 16 | 71 | 28 | 26 | 46 | 4 | 10 | 87 |
| Fayetteville | 172 | 44 | 47 | 44 | 42 | 40 | 13 | 13 | 74 | 4 | 5 | 91 | 14 | 16 | 70 | 26 | 37 | 37 | 2 | 2 | 97 |
| Greensboro | 167 | 40 | 46 | 45 | 47 | 47 | 9 | 8 | 83 | 8 | 5 | 87 | 10 | 13 | 77 | 23 | 27 | 50 | 2 | 3 | 95 |
| Greenville | 217 | 47 | 48 | 49 | 47 | 43 | 18 | 16 | 66 | 6 | 6 | 88 | 18 | 21 | 61 | 29 | 36 | 35 | 7 | 11 | 82 |
| Jacksonville | 95 | 44 | 45 | 48 | 45 | 45 | 0 | 1 | 99 | 6 | 15 | 79 | 15 | 25 | 60 | 36 | 44 | 20 | 4 | 3 | 93 |
| Laurinburg | 160 | 45 | 48 | 45 | 48 | 51 | 11 | 10 | 79 | 4 | 4 | 92 | 15 | 15 | 70 | 29 | 35 | 36 | 4 | 4 | 92 |
| Marion | 165 | 47 | 48 | 49 | 42 | 49 | 12 | 15 | 73 | 4 | 7 | 90 | 9 | 15 | 76 | 30 | 33 | 37 | 4 | 4 | 93 |
| New Hanover Co. | 209 | 44 | 55 | 50 | 45 | 44 | 16 | 6 | 78 | 3 | 2 | 95 | 14 | 15 | 71 | 25 | 27 | 48 | 7 | 2 | 90 |
| Raleigh 1 | 207 | 48 | 52 | 52 | 51 | 48 | 18 | 14 | 68 | 9 | 6 | 85 | 13 | 21 | 66 | 33 | 33 | 34 | 5 | 5 | 89 |
| Raleigh 2 | 138 | 41 | 46 | 46 | 46 | 45 | 9 | 14 | 78 | 1 | 12 | 88 | 13 | 20 | 67 | 24 | 36 | 40 | 2 | 4 | 94 |
| Raleigh 3 | 139 | 41 | 42 | 47 | 49 | 46 | 10 | 11 | 79 | 7 | 4 | 88 | 11 | 17 | 73 | 27 | 31 | 42 | 4 | 4 | 93 |
| Roanoke Rapids | 170 | 38 | 49 | 44 | 45 | 41 | 12 | 11 | 77 | 4 | 4 | 93 | 8 | 15 | 77 | 18 | 29 | 53 | 0 | 4 | 96 |
| Tuckaseigee | 200 | 50 | 53 | 50 | 56 | 52 | 12 | 10 | 77 | 7 | 6 | 87 | 14 | 10 | 76 | 35 | 26 | 38 | 11 | 6 | 83 |
| Wilmington | 213 | 43 | 50 | 49 | 44 | 43 | 15 | 9 | 75 | 4 | 4 | 92 | 15 | 17 | 67 | 32 | 36 | 32 | 5 | 15 | 81 |
| Wilson | 169 | 41 | 45 | 45 | 44 | 43 | 9 | 14 | 77 | 7 | 2 | 91 | 14 | 24 | 62 | 24 | 36 | 41 | 0 | 3 | 97 |
| Winston-Salem | 163 | 50 | 48 | 47 | 52 | 50 | 11 | 12 | 77 | 6 | 4 | 90 | 10 | 20 | 70 | 31 | 32 | 37 | 4 | 5 | 91 |

[a] Out of three CDC classes: increasing (↑), decreasing (↓), and plateau (—)

### S13 Sewershed-Specific Rate of Change Estimates

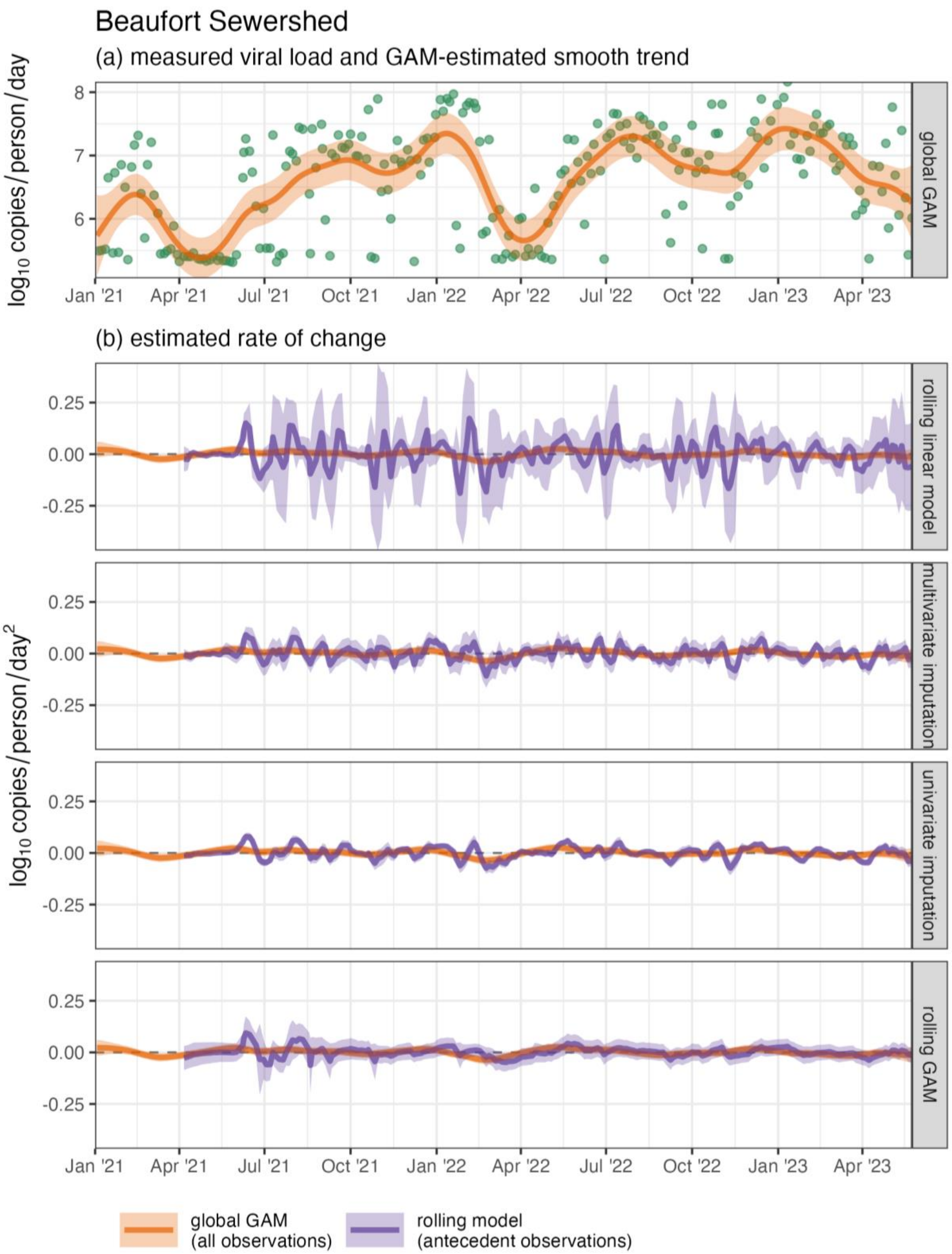

Figure S7. Measured SARS-CoV-2 wastewater per-capita viral load (green points) and estimated mean and 95% CI of the (a) temporal trend and (b) rates of change in the trend for the Beaufort sewershed

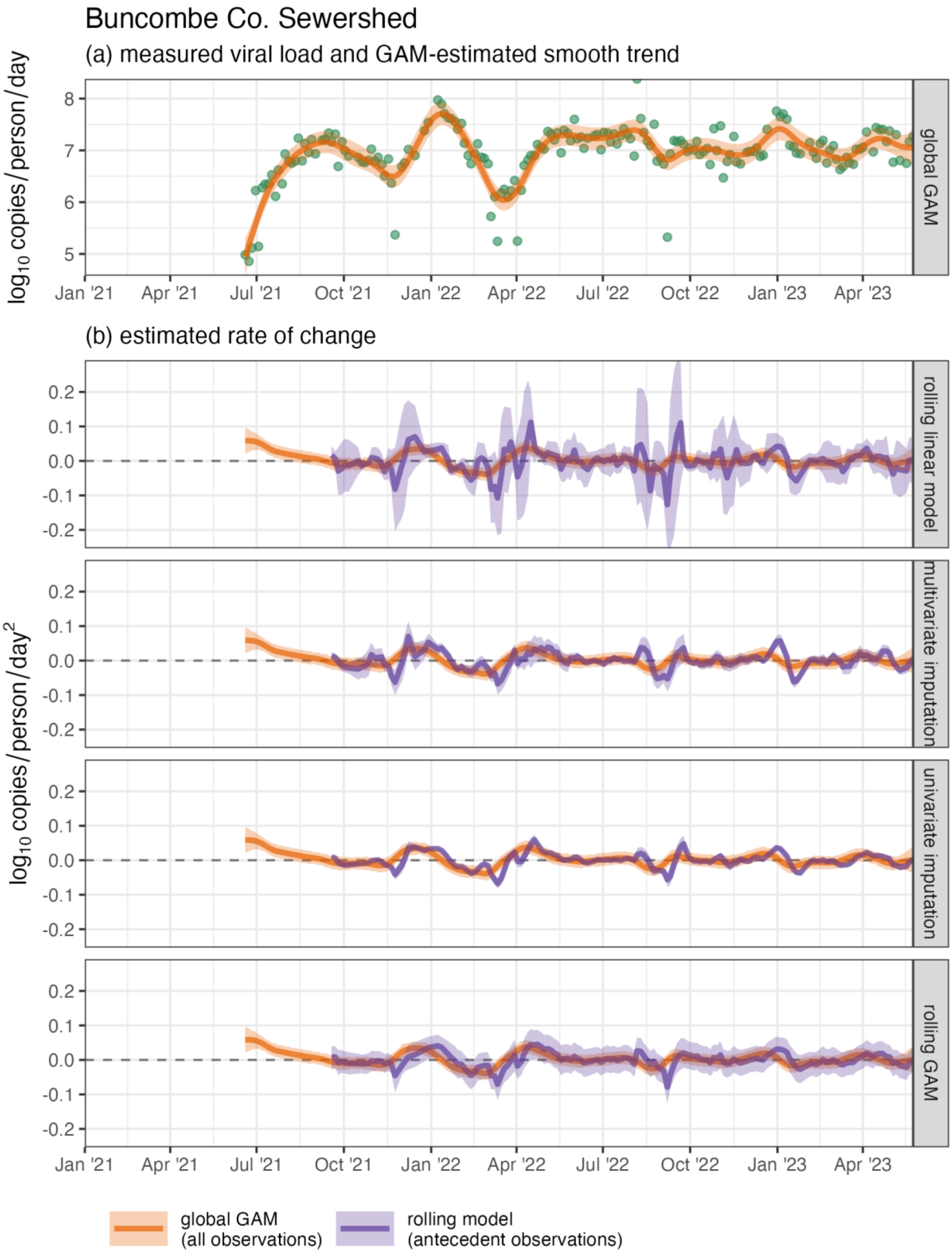

Figure S8. Measured SARS-CoV-2 wastewater per-capita viral load (green points) and estimated mean and 95% CI of the (a) temporal trend and (b) rates of change in the trend for the Buncombe County sewershed

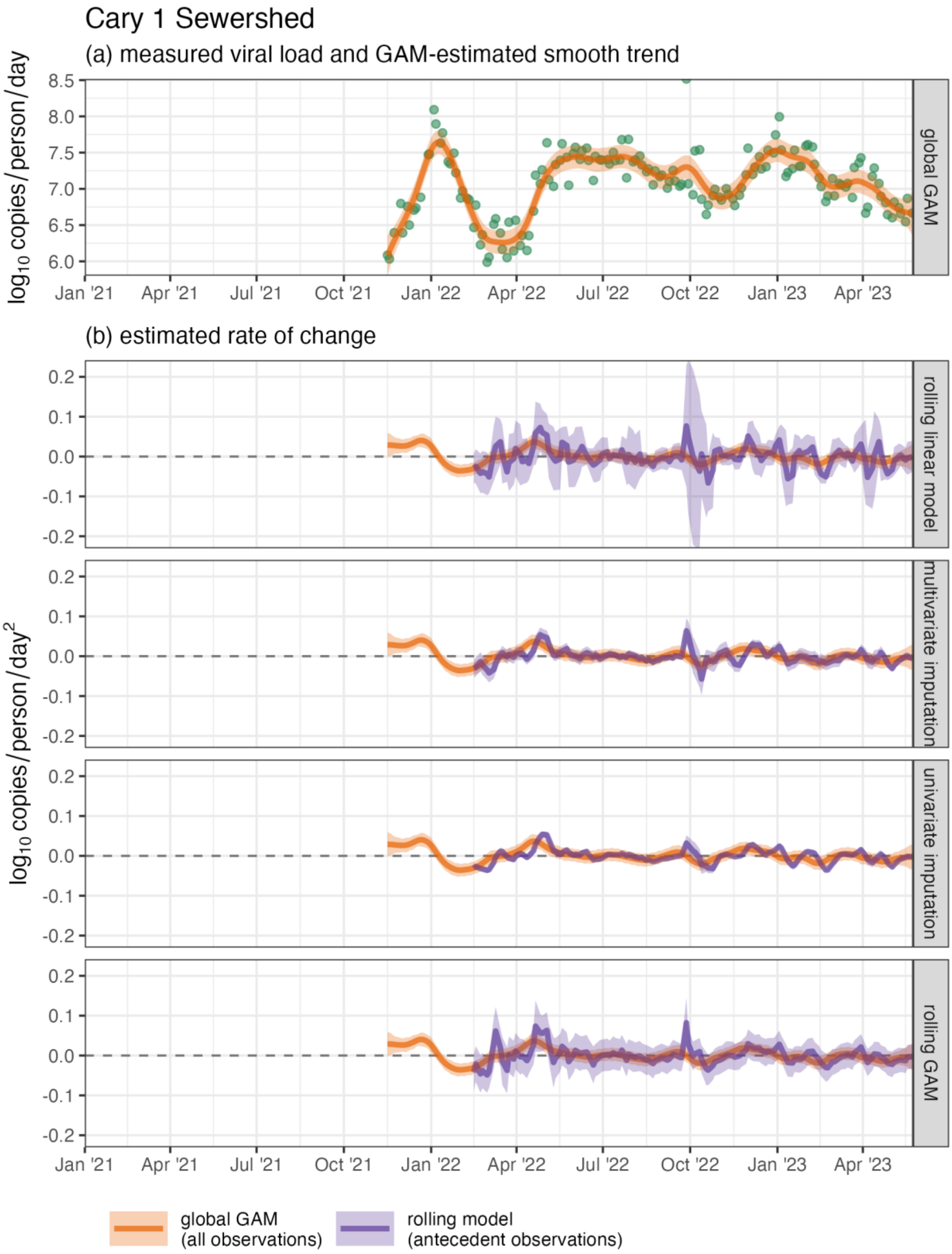

Figure S9. Measured SARS-CoV-2 wastewater per-capita viral load (green points) and estimated mean and 95% CI of the (a) temporal trend and (b) rates of change in the trend for the Cary 1 sewershed

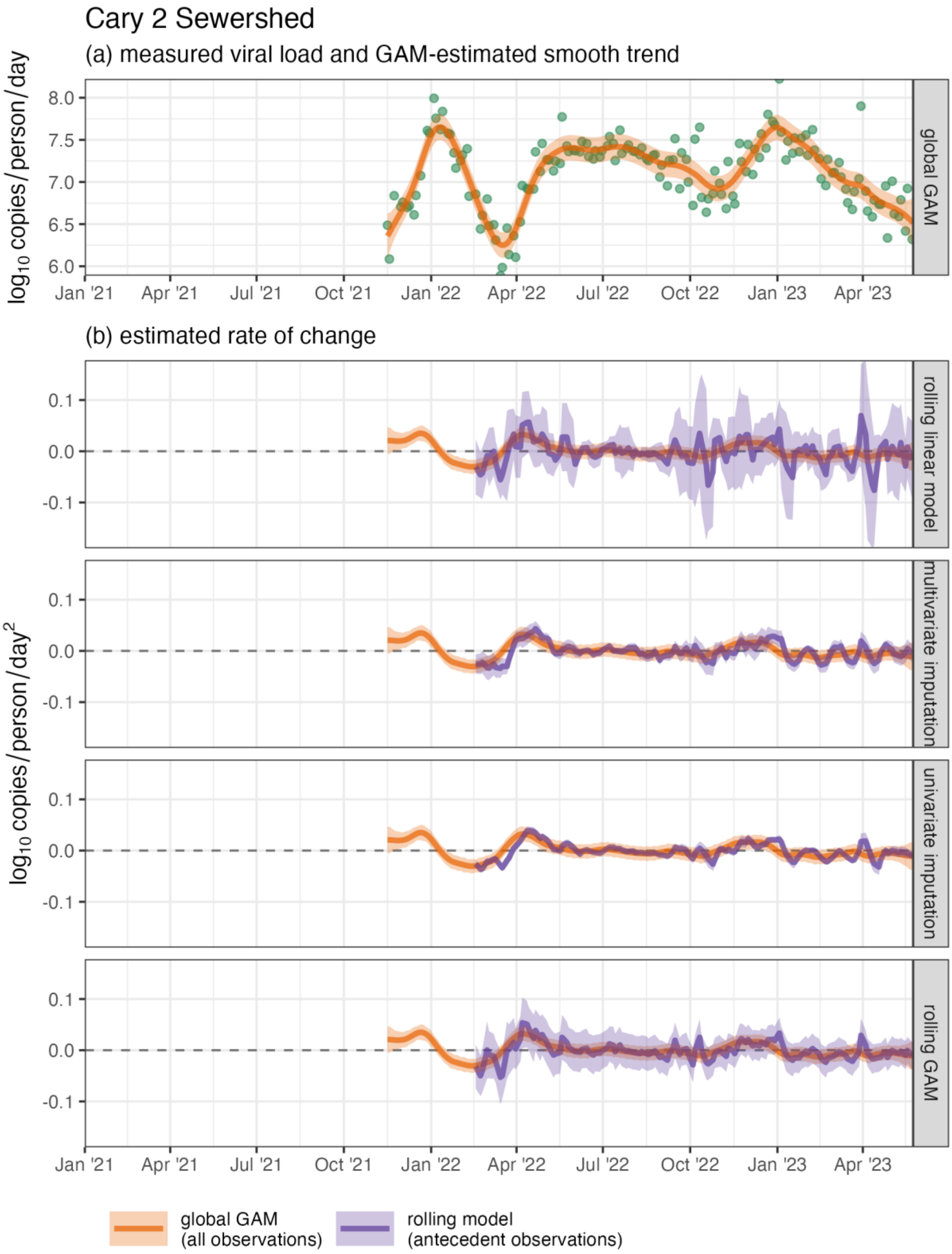

Figure S10. Measured SARS-CoV-2 wastewater per-capita viral load (green points) and estimated mean and 95% CI of the (a) temporal trend and (b) rates of change in the trend for the Cary 2 sewershed

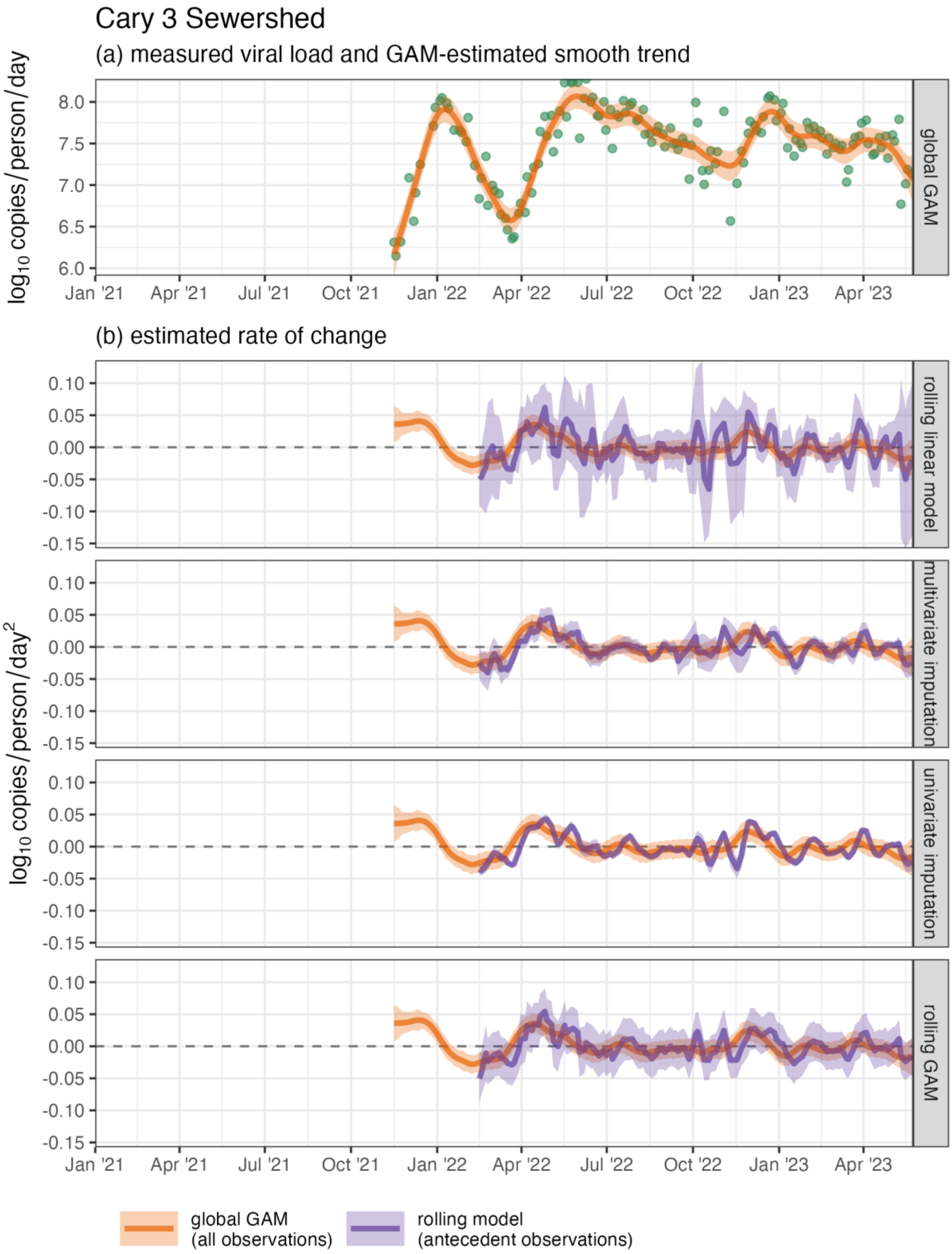

Figure S11. Measured SARS-CoV-2 wastewater per-capita viral load (green points) and estimated mean and 95% CI of the (a) temporal trend and (b) rates of change in the trend for the Cary 3 sewershed

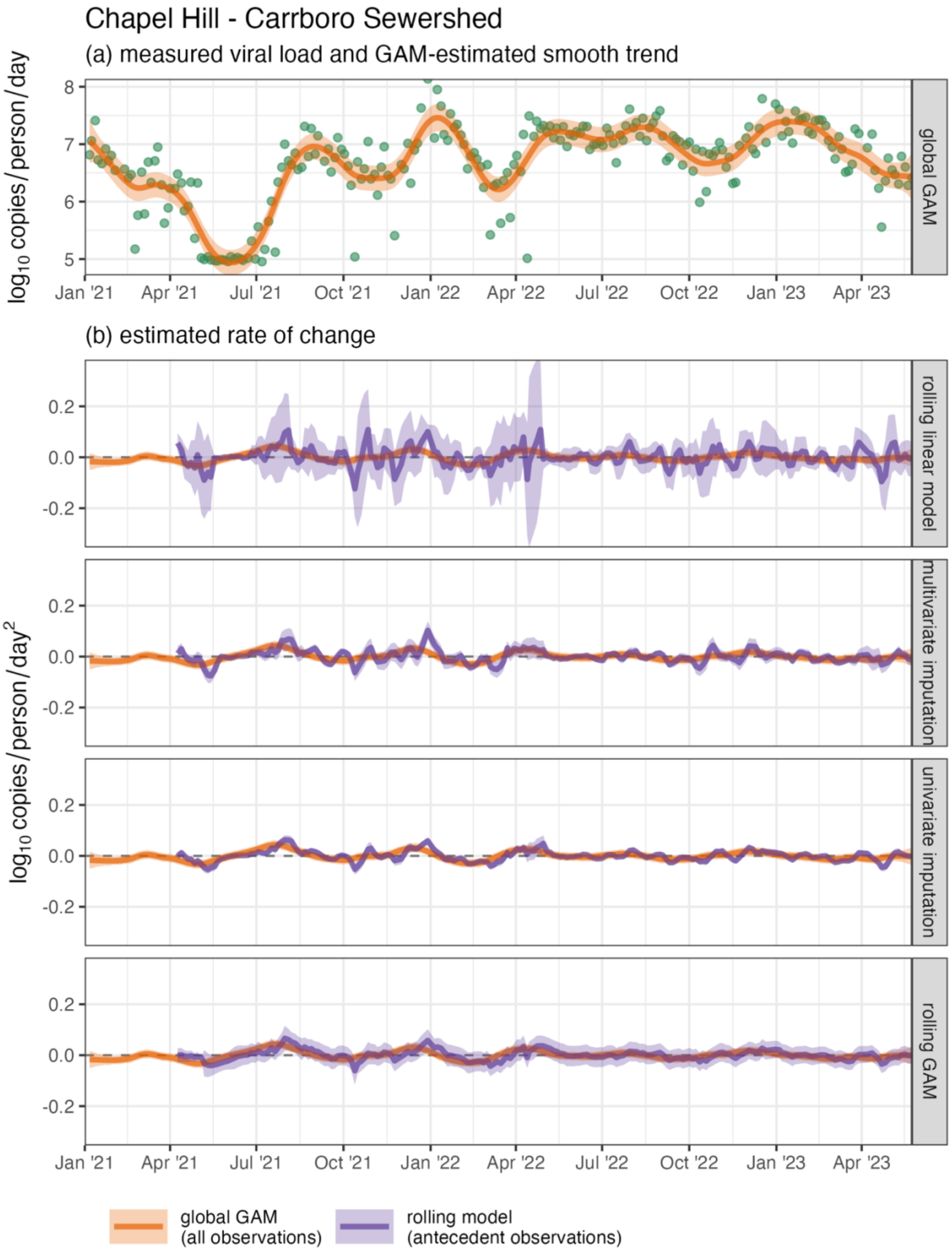

Figure S12. Measured SARS-CoV-2 wastewater per-capita viral load (green points) and estimated mean and 95% CI of the (a) temporal trend and (b) rates of change in the trend for the Chapel Hill – Carrboro sewershed

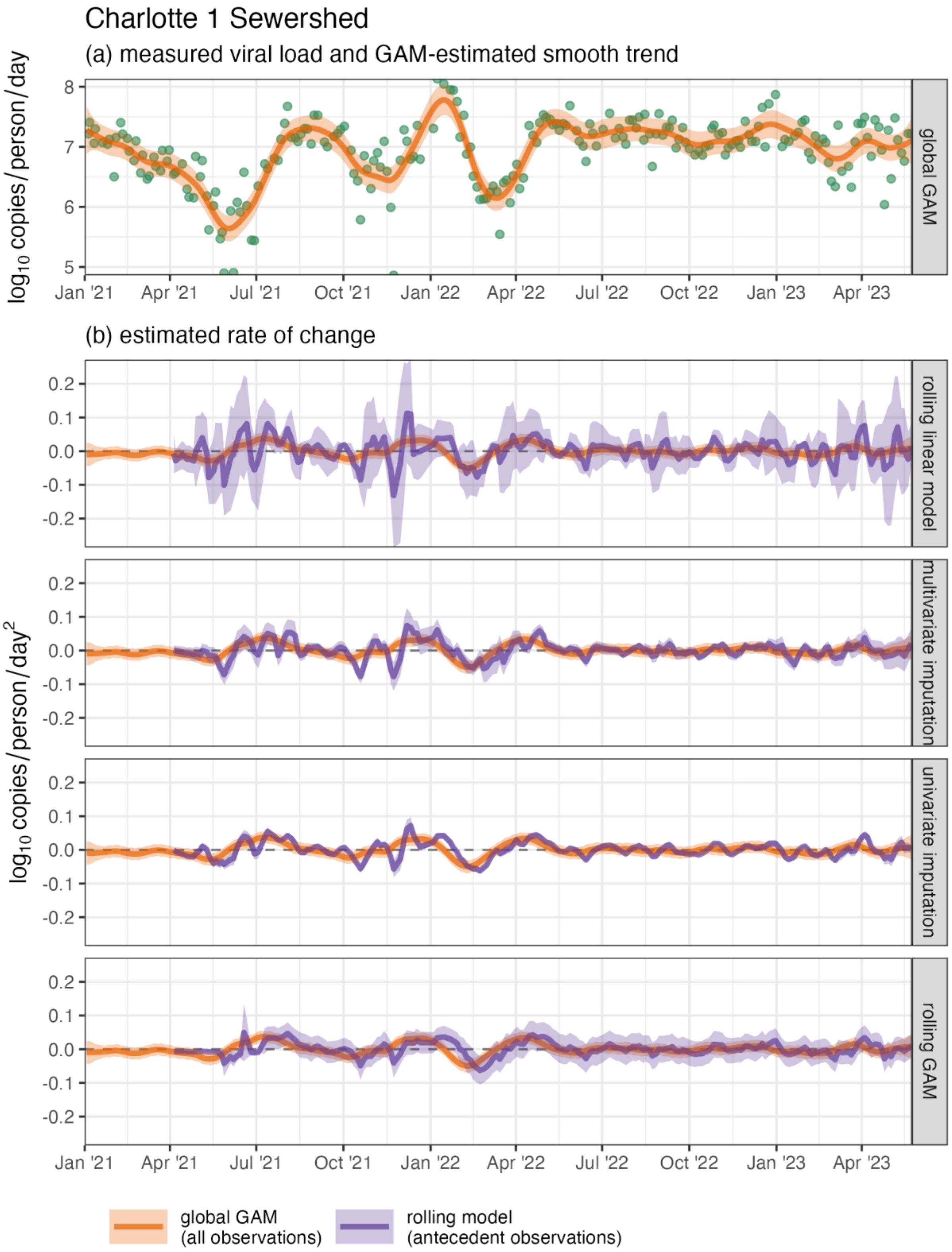

Figure S13. Measured SARS-CoV-2 wastewater per-capita viral load (green points) and estimated mean and 95% CI of the (a) temporal trend and (b) rates of change in the trend for the Charlotte 1 sewershed

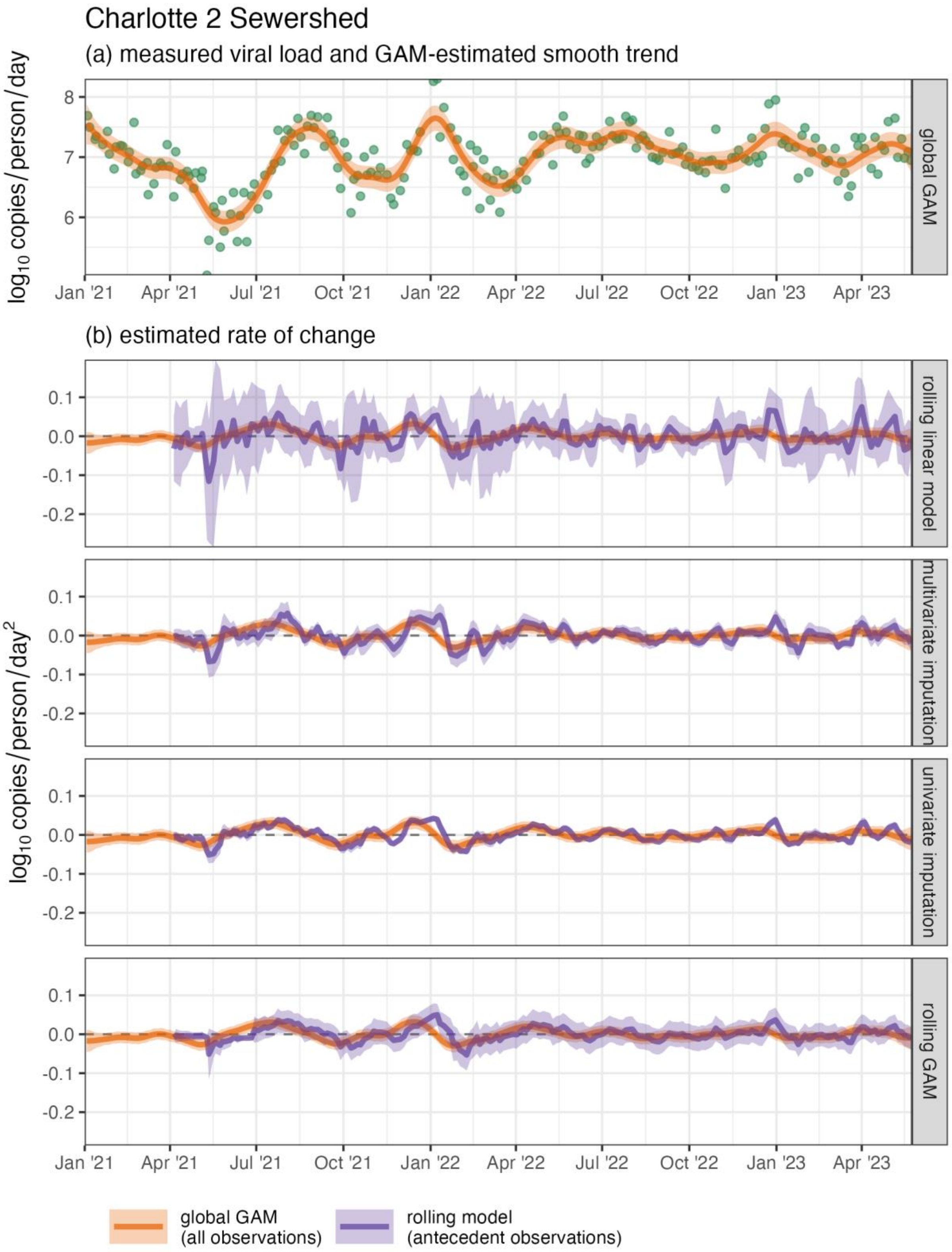

Figure S14. Measured SARS-CoV-2 wastewater per-capita viral load (green points) and estimated mean and 95% CI of the (a) temporal trend and (b) rates of change in the trend for the Charlotte 2 sewershed

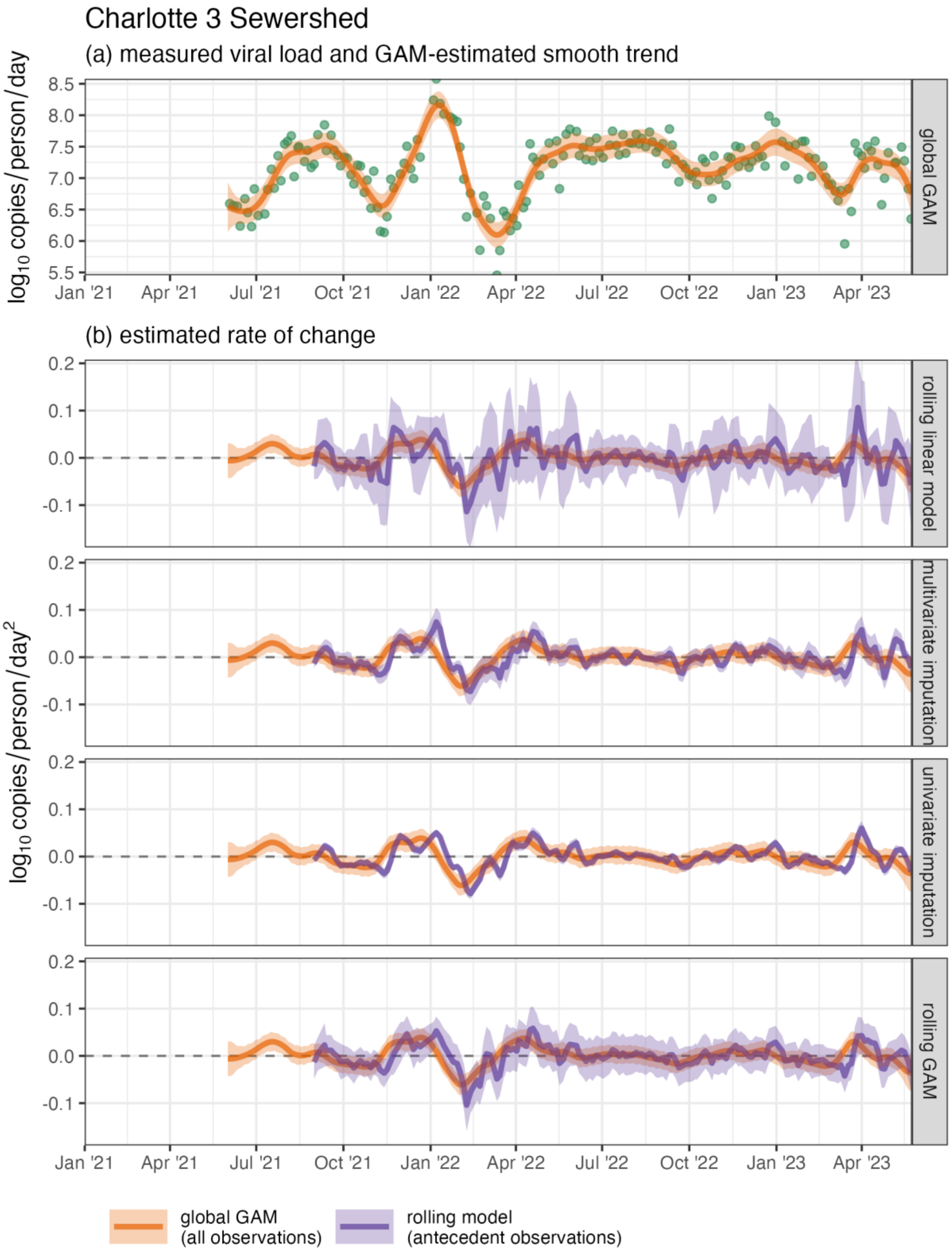

Figure S15. Measured SARS-CoV-2 wastewater per-capita viral load (green points) and estimated mean and 95% CI of the (a) temporal trend and (b) rates of change in the trend for the Charlotte 3 sewershed

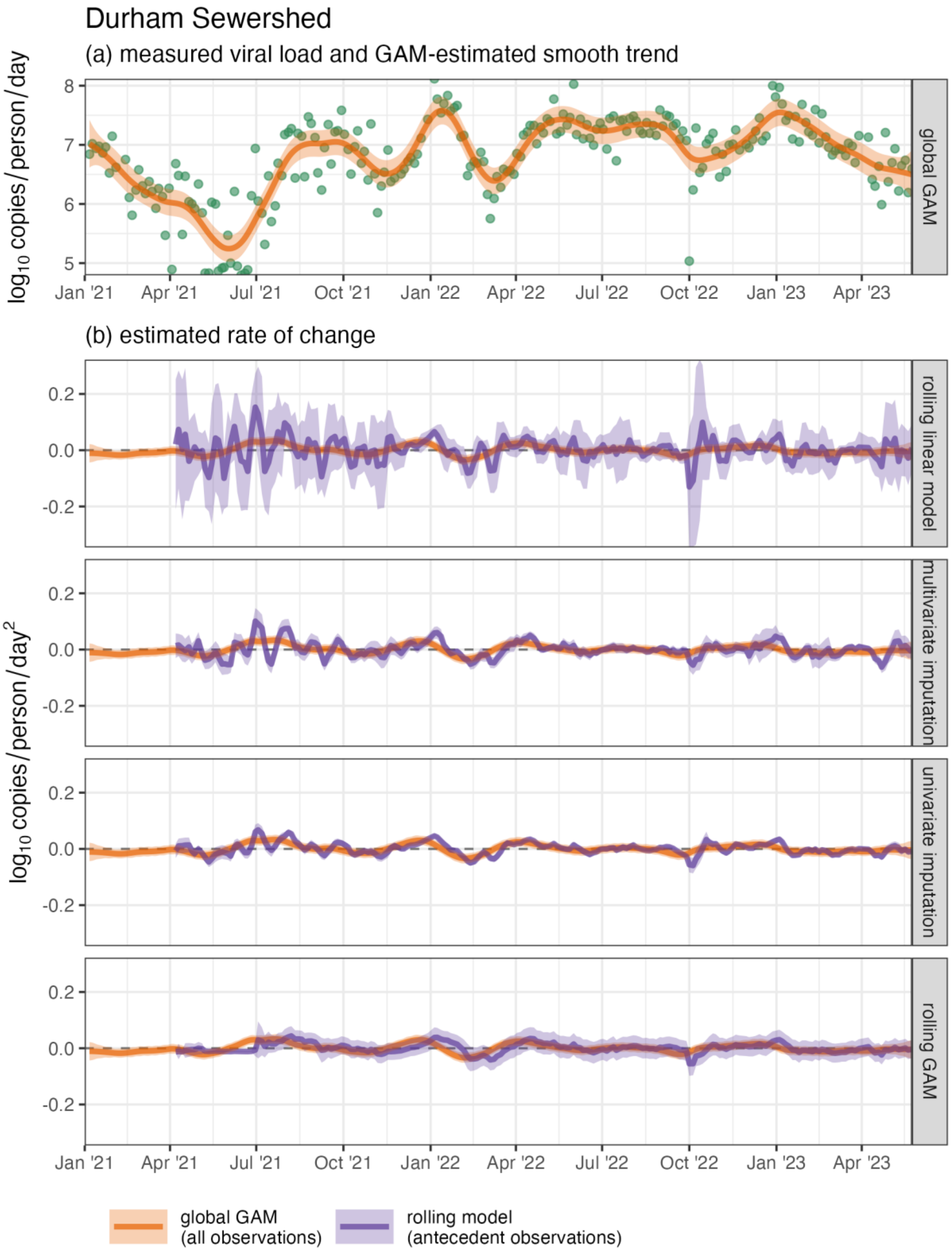

Figure S16. Measured SARS-CoV-2 wastewater per-capita viral load (green points) and estimated mean and 95% CI of the (a) temporal trend and (b) rates of change in the trend for the Durham sewershed

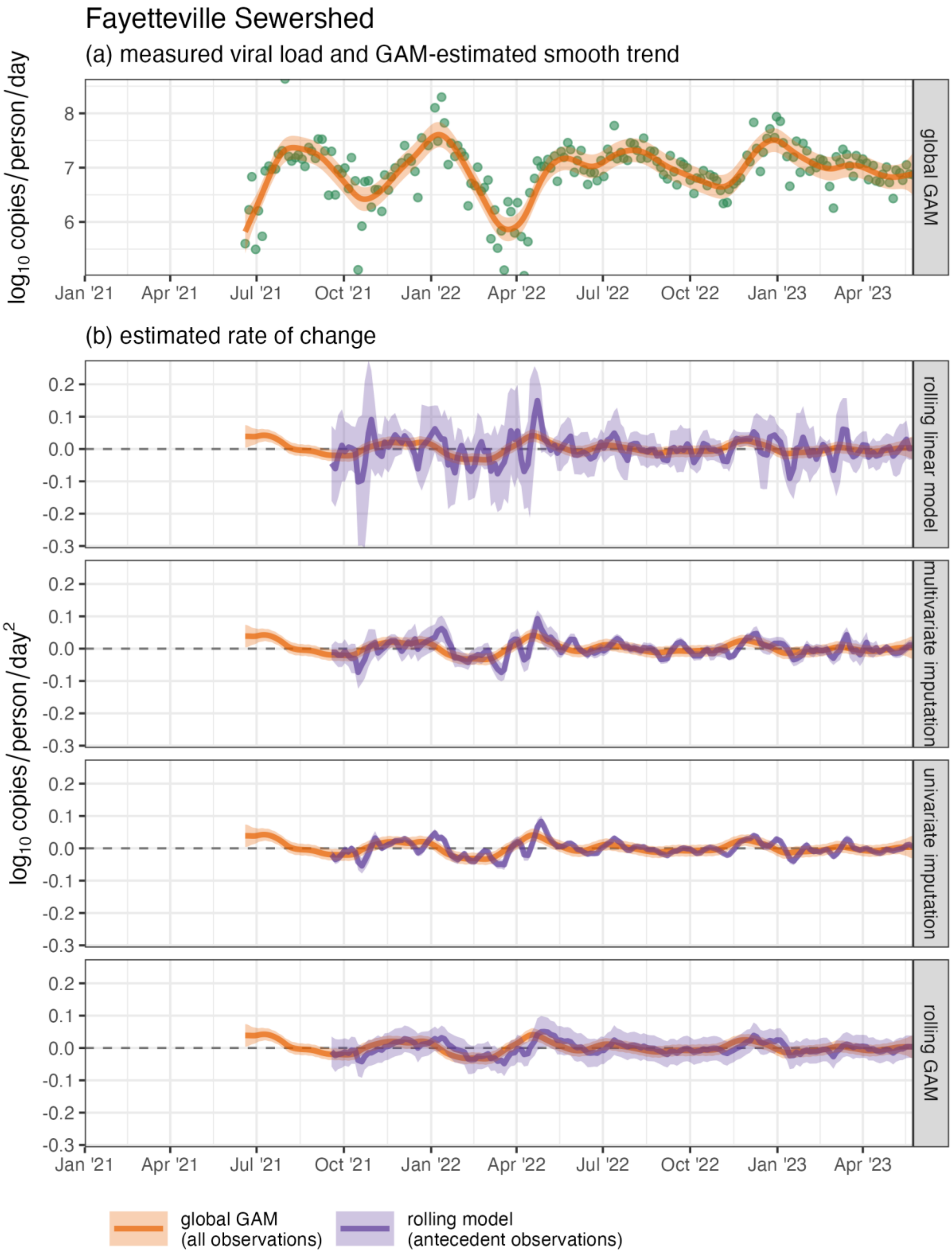

Figure S17. Measured SARS-CoV-2 wastewater per-capita viral load (green points) and estimated mean and 95% CI of the (a) temporal trend and (b) rates of change in the trend for the Fayetteville sewershed

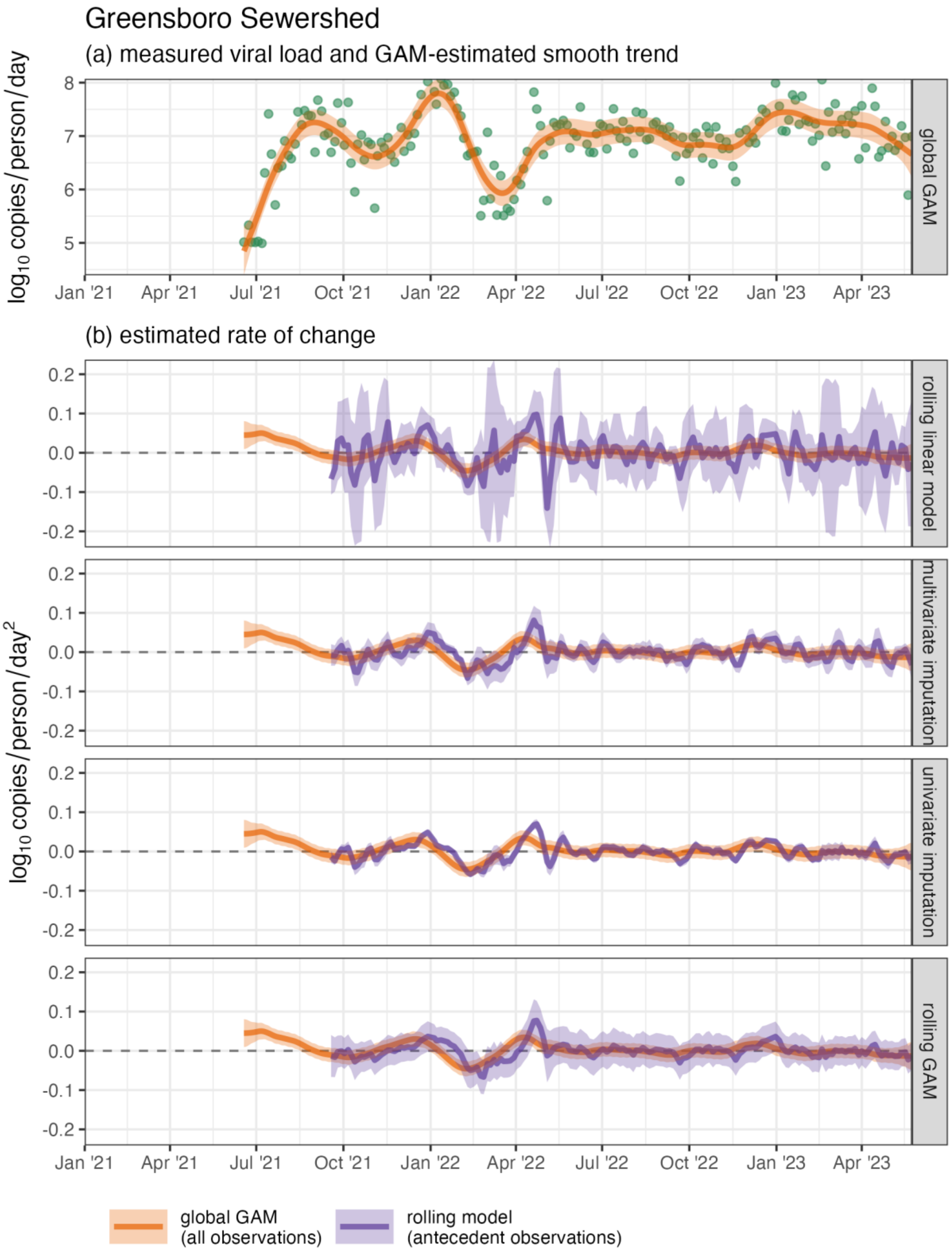

Figure S18. Measured SARS-CoV-2 wastewater per-capita viral load (green points) and estimated mean and 95% CI of the (a) temporal trend and (b) rates of change in the trend for the Greensboro sewershed

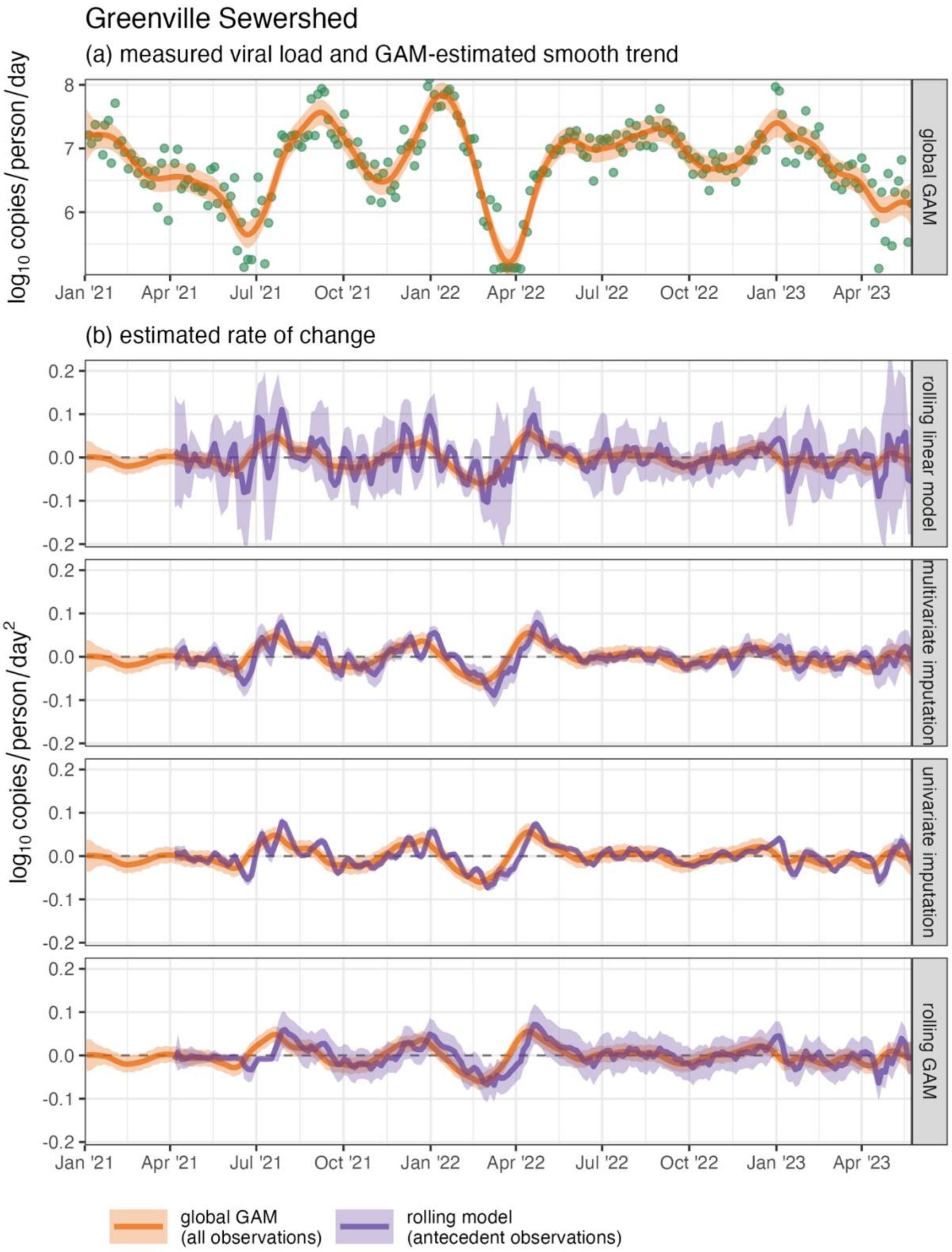

Figure S19. Measured SARS-CoV-2 wastewater per-capita viral load (green points) and estimated mean and 95% CI of the (a) temporal trend and (b) rates of change in the trend for the Greenville sewershed

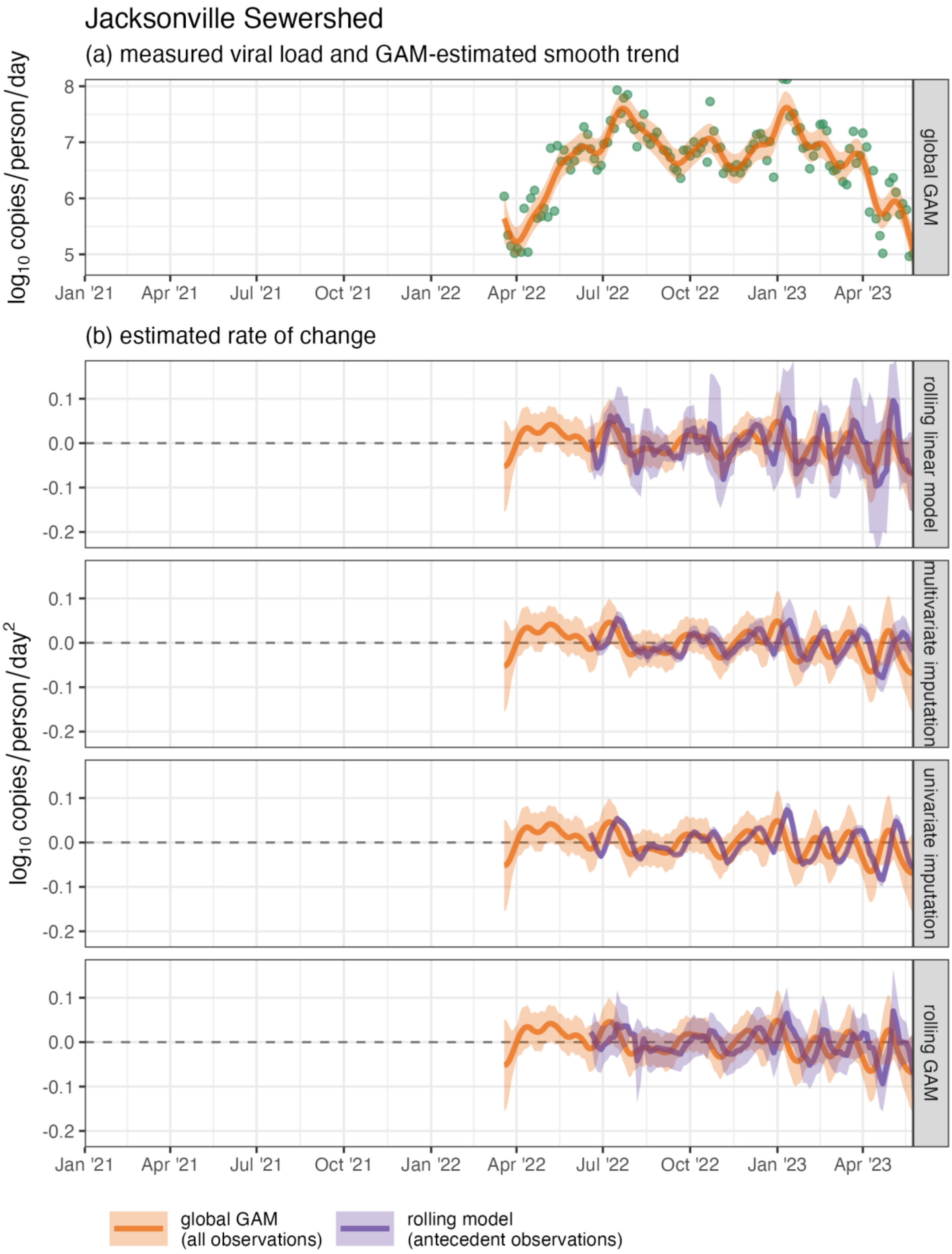

Figure S20. Measured SARS-CoV-2 wastewater per-capita viral load (green points) and estimated mean and 95% CI of the (a) temporal trend and (b) rates of change in the trend for the Jacksonville sewershed

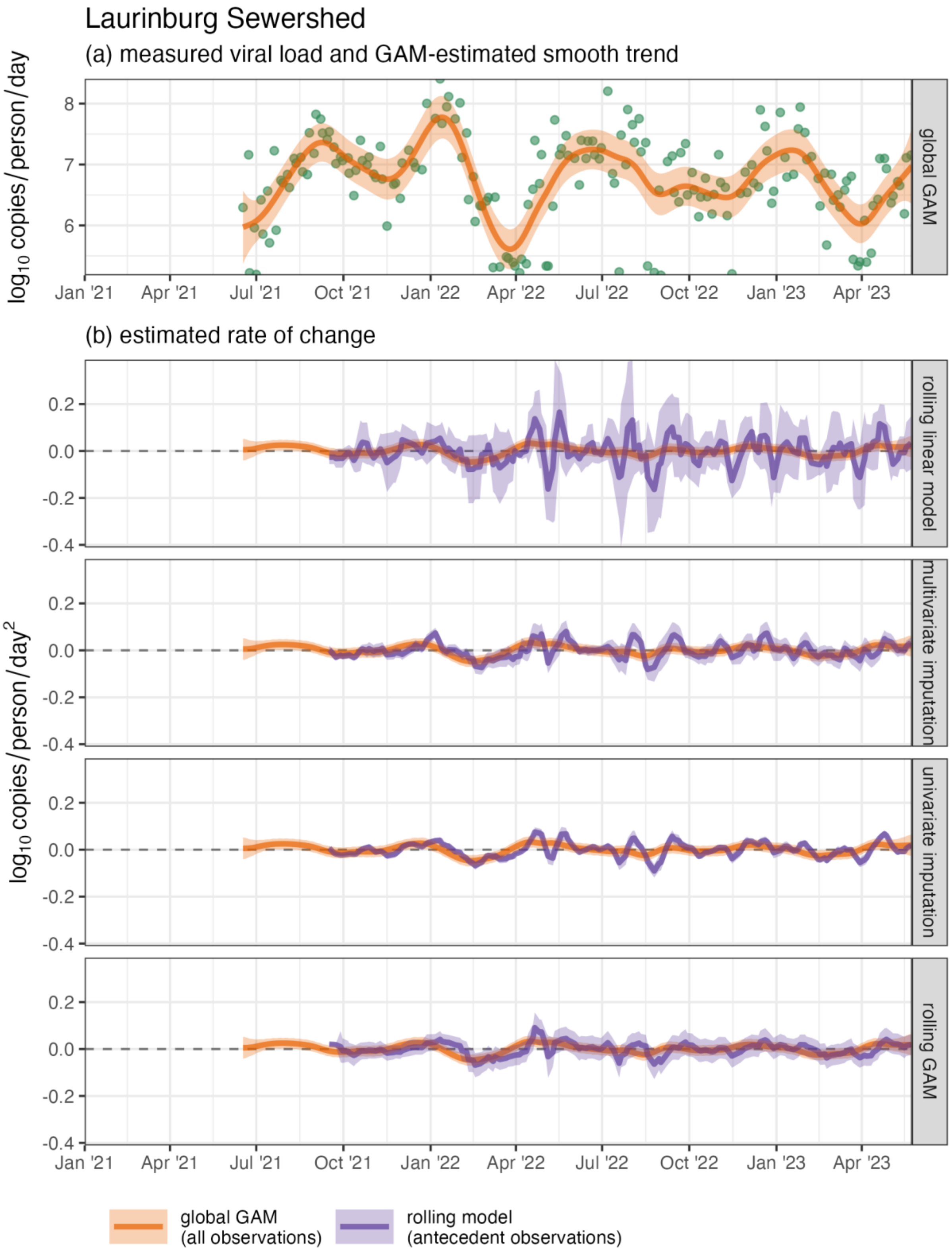

Figure S21. Measured SARS-CoV-2 wastewater per-capita viral load (green points) and estimated mean and 95% CI of the (a) temporal trend and (b) rates of change in the trend for the Laurinburg sewershed

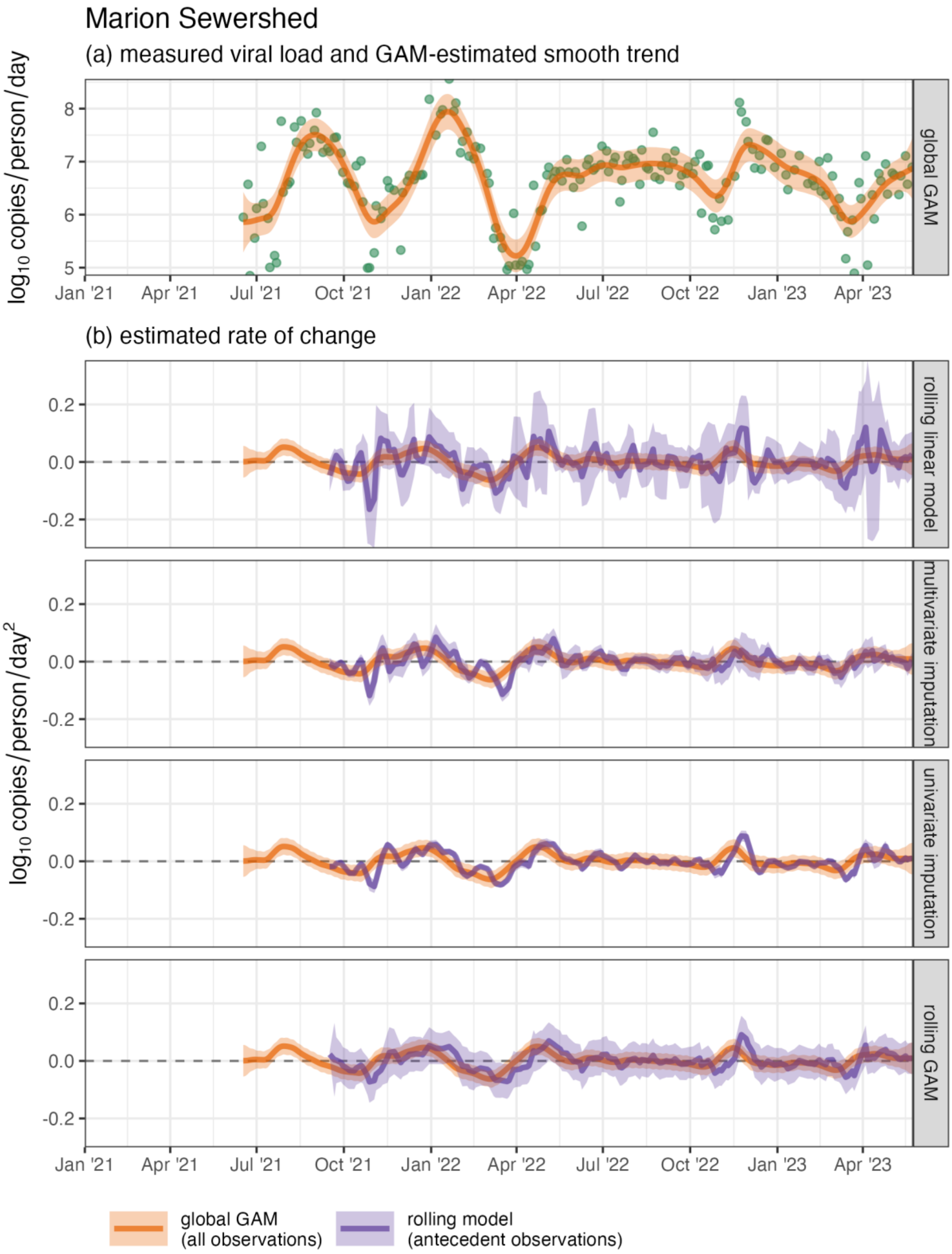

Figure S22. Measured SARS-CoV-2 wastewater per-capita viral load (green points) and estimated mean and 95% CI of the (a) temporal trend and (b) rates of change in the trend for the Marion sewershed

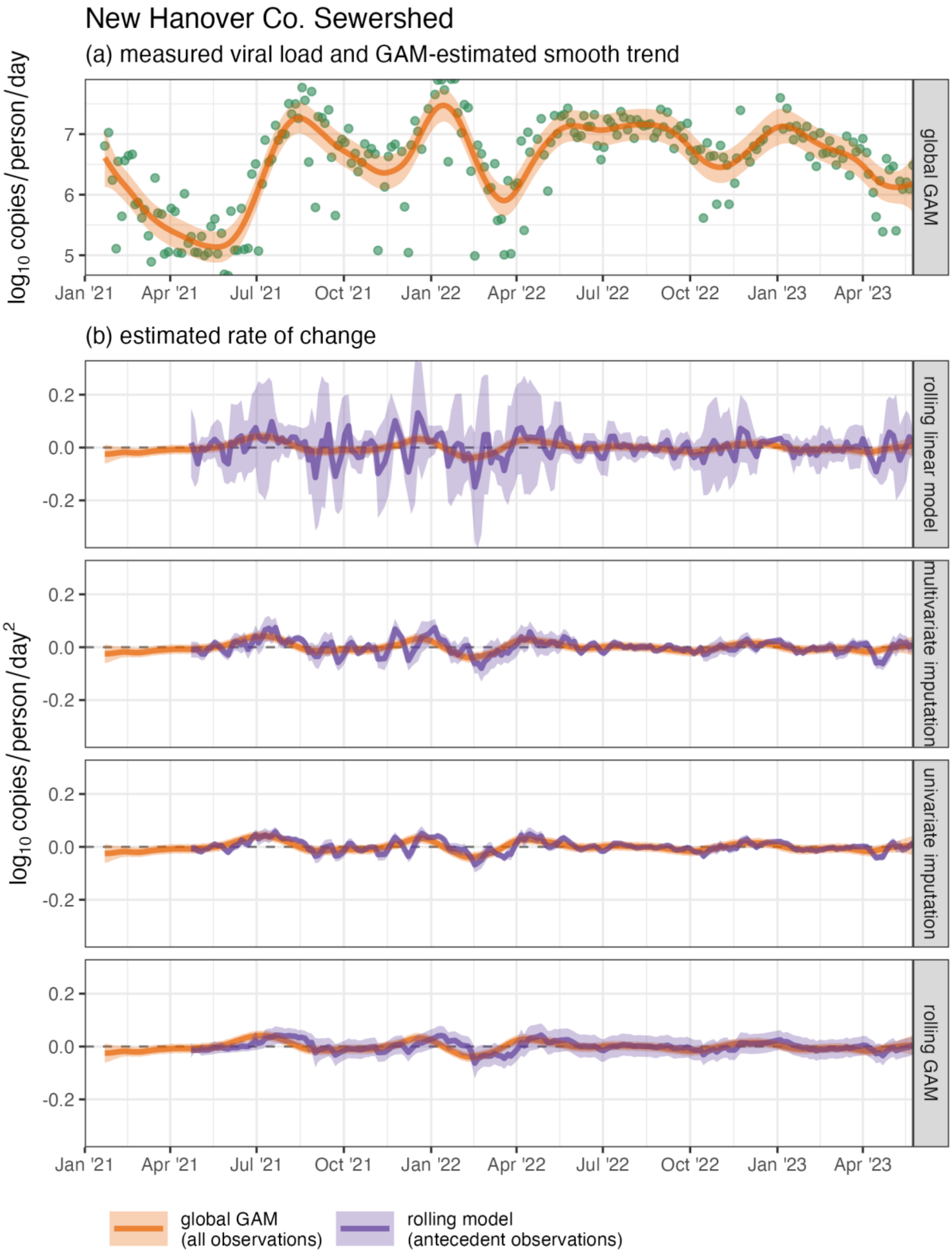

Figure S23. Measured SARS-CoV-2 wastewater per-capita viral load (green points) and estimated mean and 95% CI of the (a) temporal trend and (b) rates of change in the trend for the New Hanover County sewershed

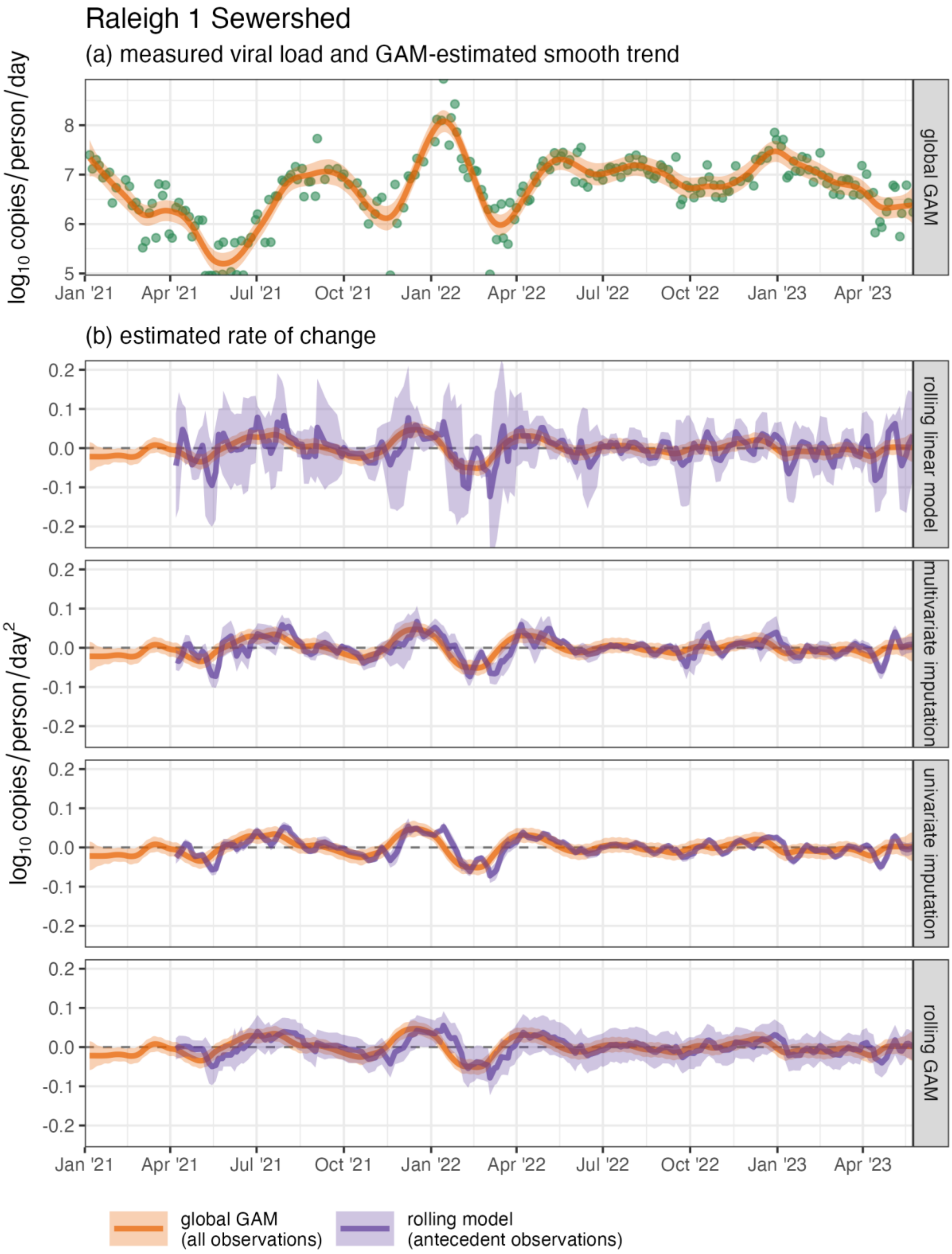

Figure S24. Measured SARS-CoV-2 wastewater per-capita viral load (green points) and estimated mean and 95% CI of the (a) temporal trend and (b) rates of change in the trend for the Raleigh 1 sewershed

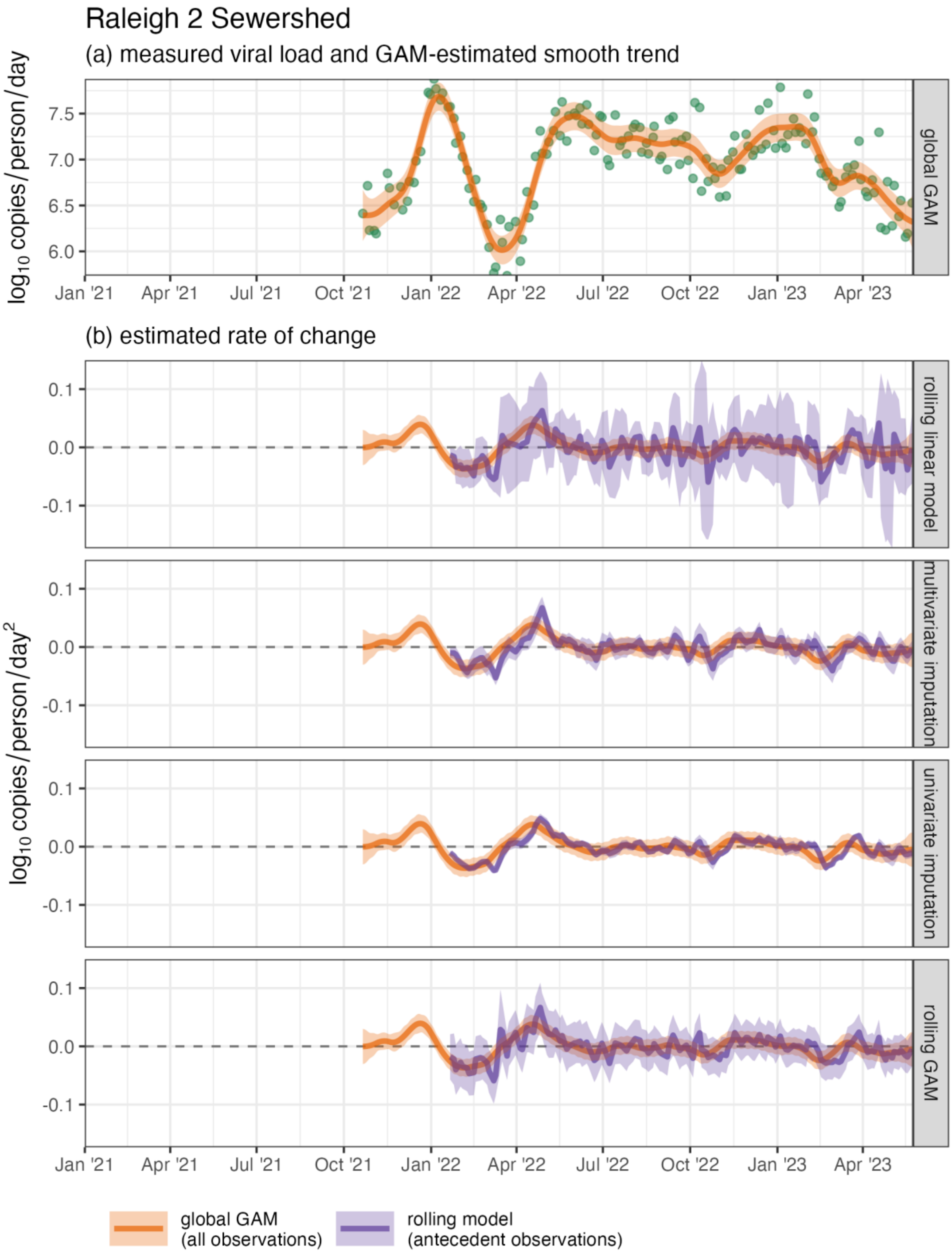

Figure S25. Measured SARS-CoV-2 wastewater per-capita viral load (green points) and estimated mean and 95% CI of the (a) temporal trend and (b) rates of change in the trend for the Raleigh 2 sewershed

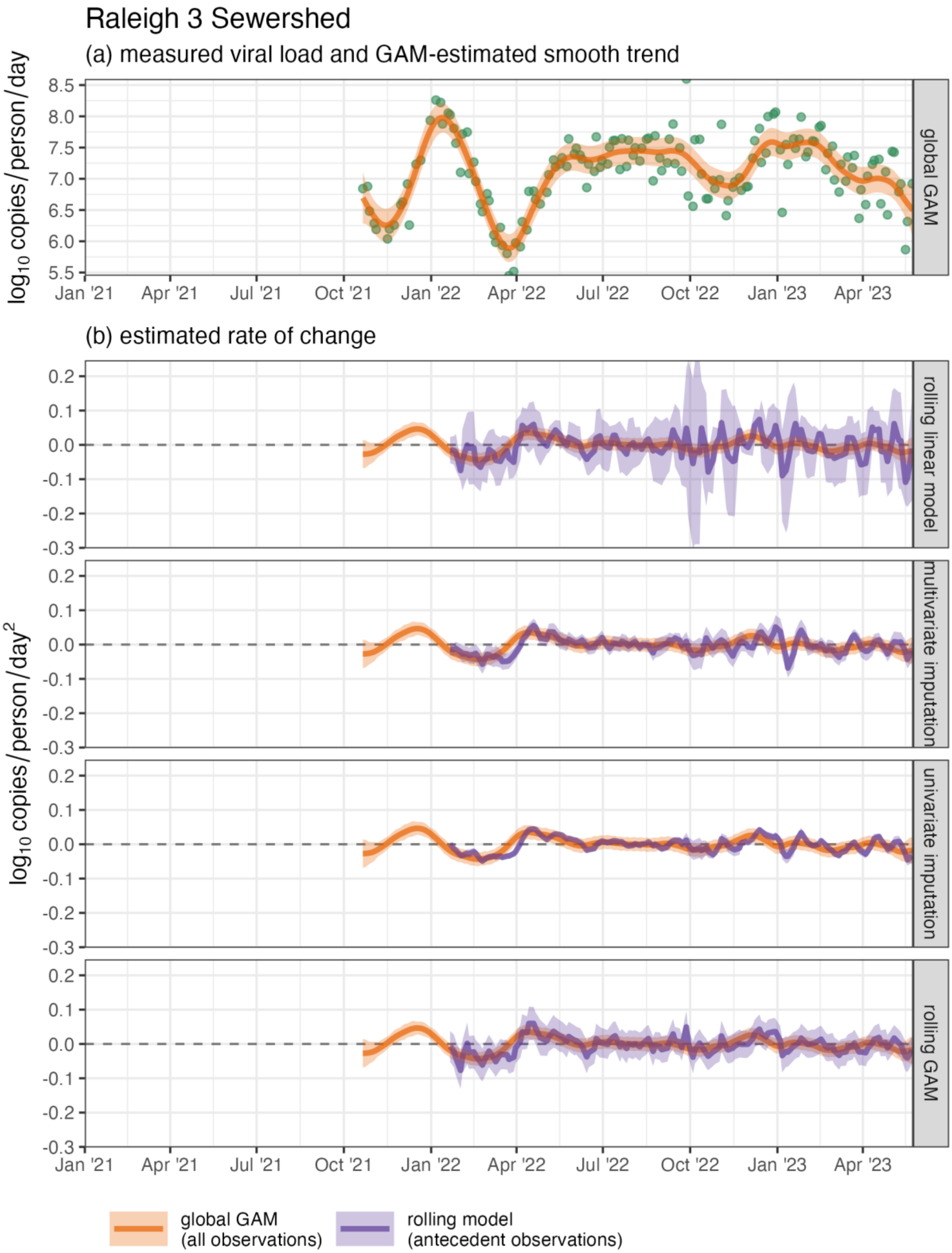

Figure S26. Measured SARS-CoV-2 wastewater per-capita viral load (green points) and estimated mean and 95% CI of the (a) temporal trend and (b) rates of change in the trend for the Raleigh 3 sewershed

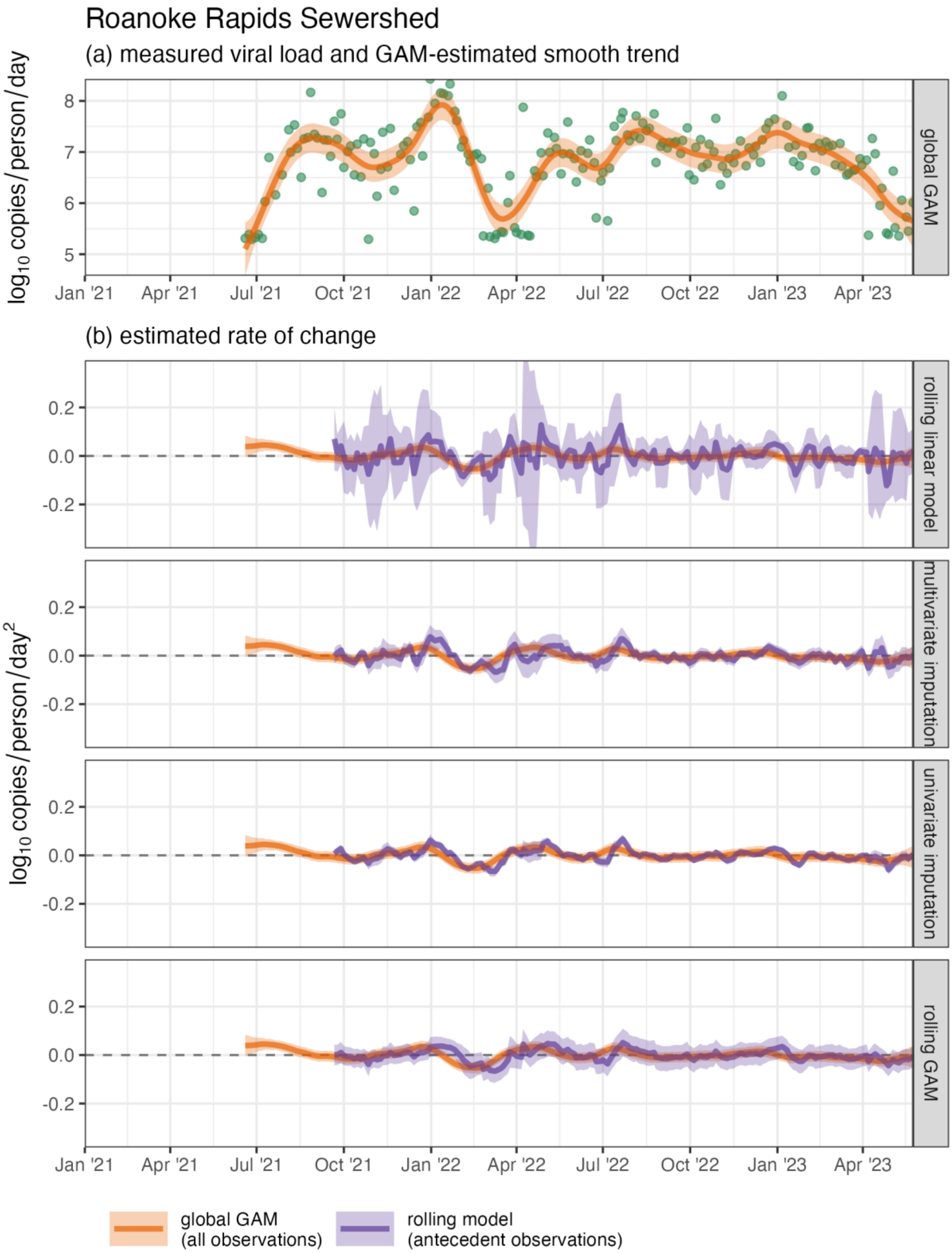

Figure S27. Measured SARS-CoV-2 wastewater per-capita viral load (green points) and estimated mean and 95% CI of the (a) temporal trend and (b) rates of change in the trend for the Roanoke Rapids sewershed

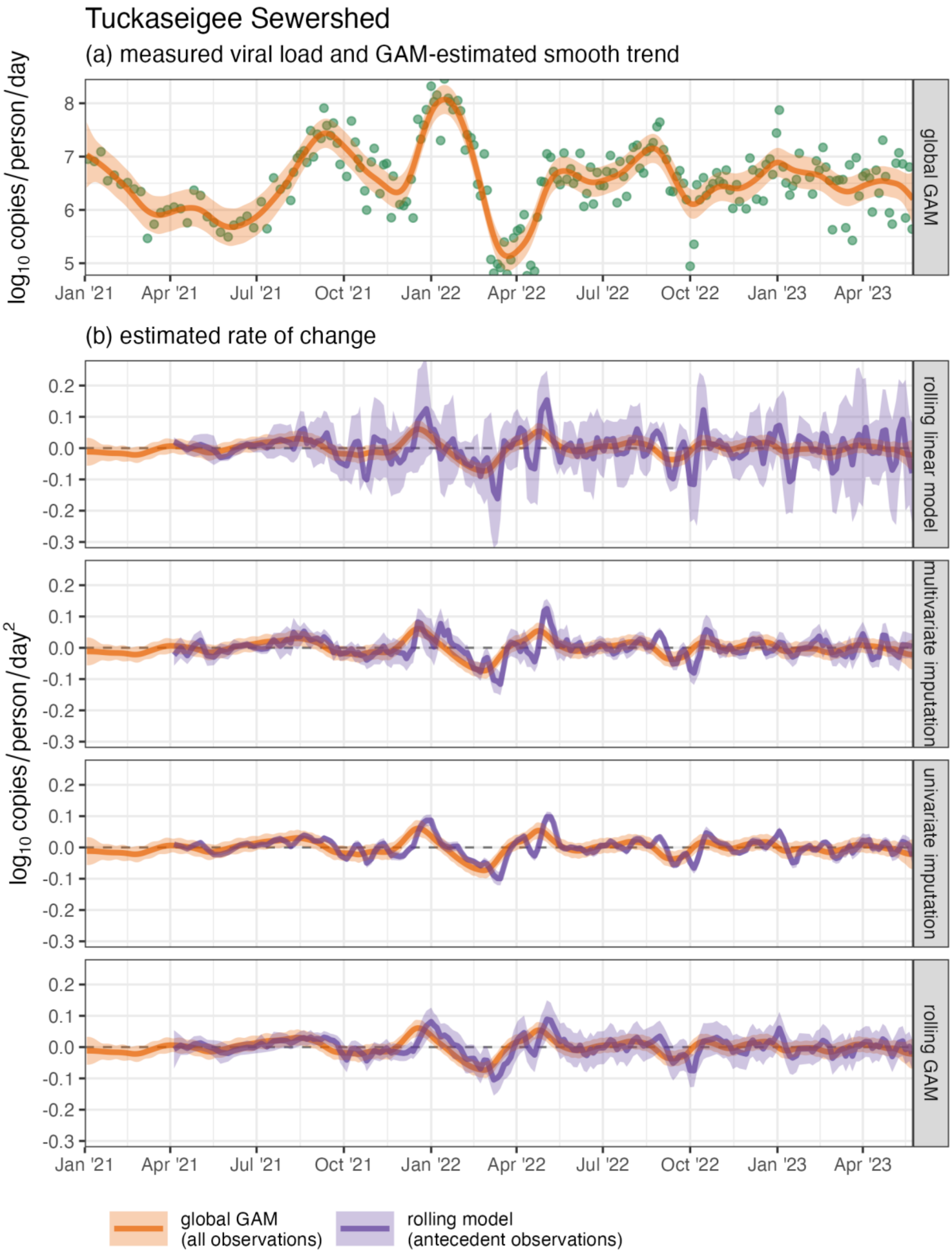

Figure S28. Measured SARS-CoV-2 wastewater per-capita viral load (green points) and estimated mean and 95% CI of the (a) temporal trend and (b) rates of change in the trend for the Tuckaseigee sewershed

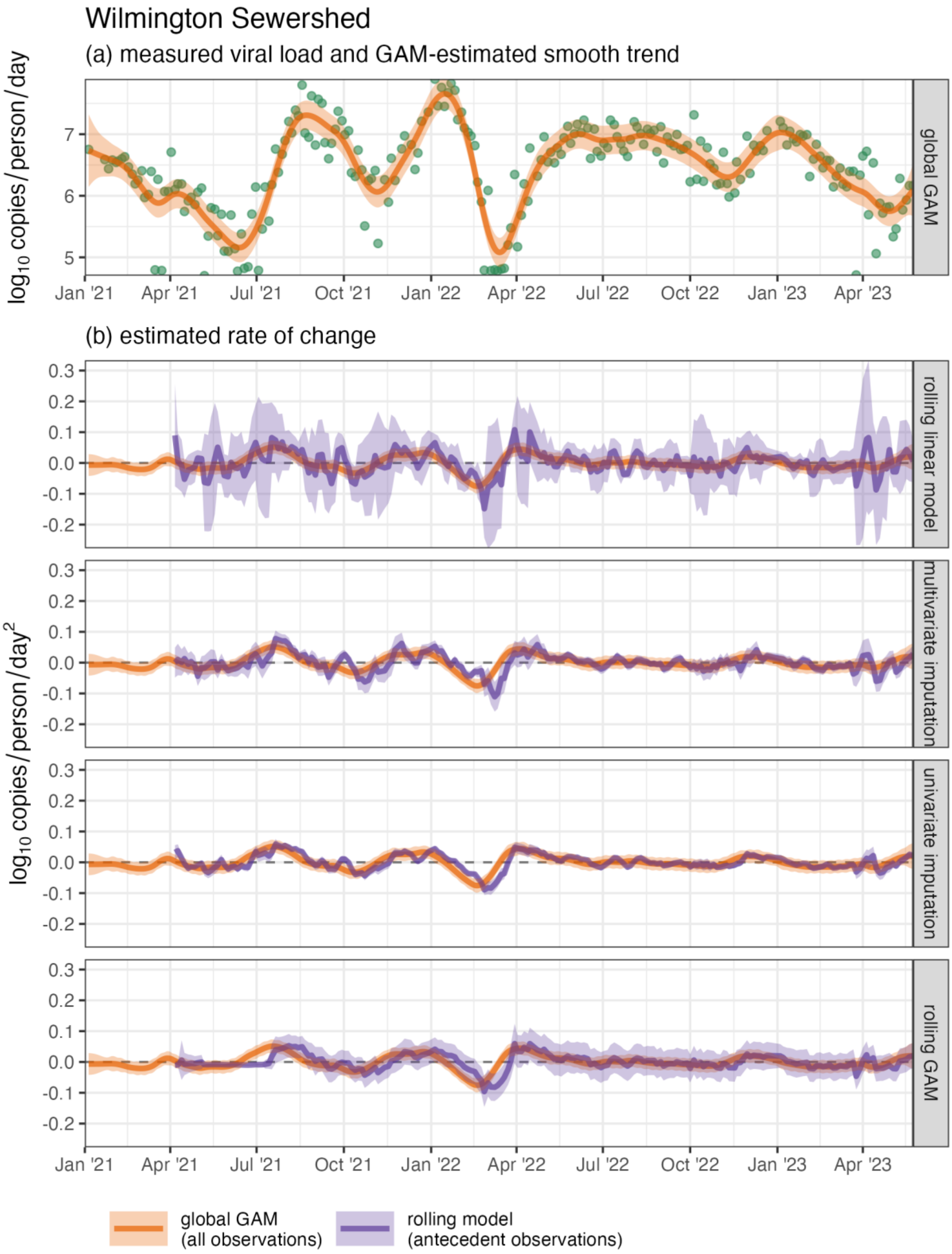

Figure S29. Measured SARS-CoV-2 wastewater per-capita viral load (green points) and estimated mean and 95% CI of the (a) temporal trend and (b) rates of change in the trend for the Wilmington sewershed

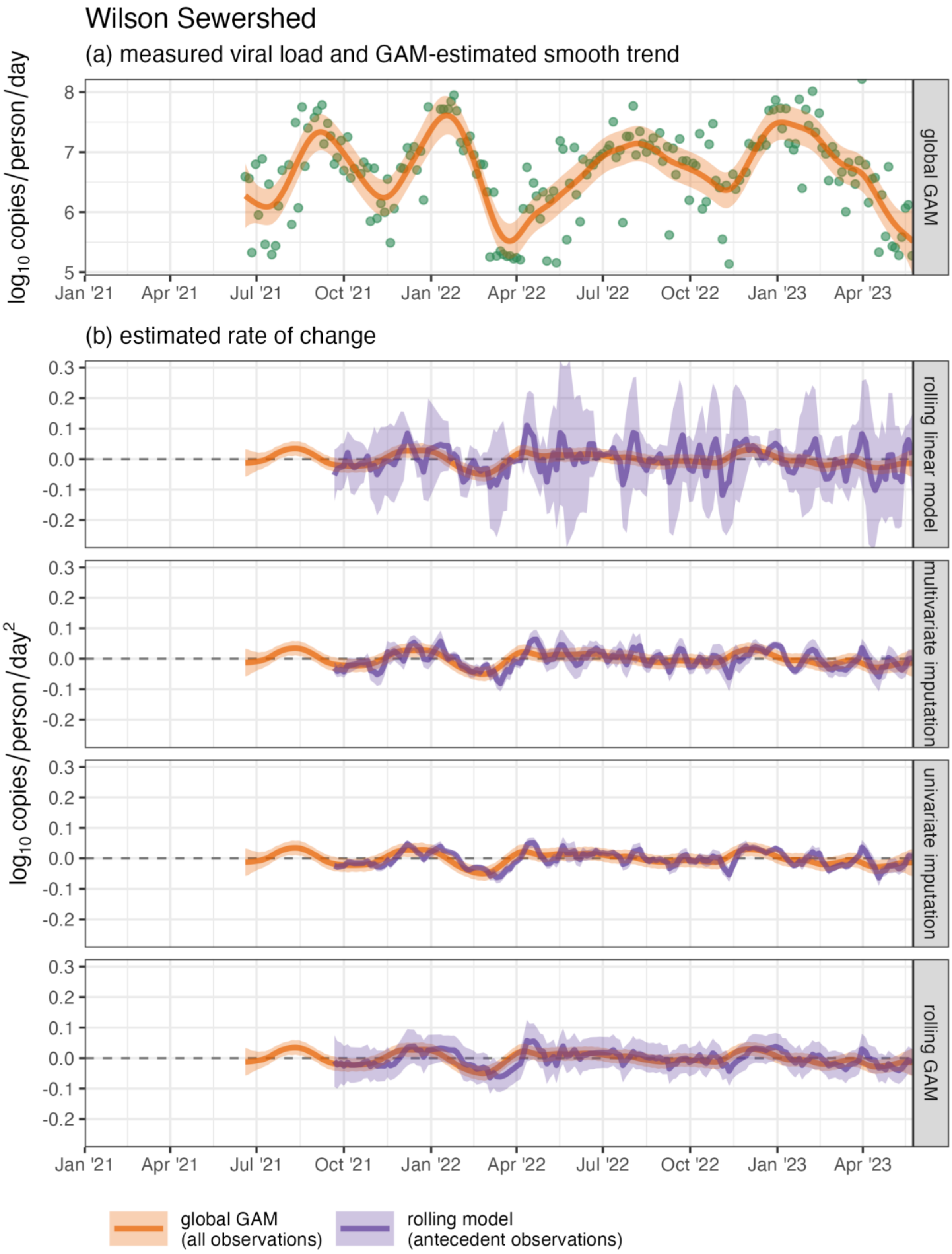

Figure S30. Measured SARS-CoV-2 wastewater per-capita viral load (green points) and estimated mean and 95% CI of the (a) temporal trend and (b) rates of change in the trend for the Wilson sewershed

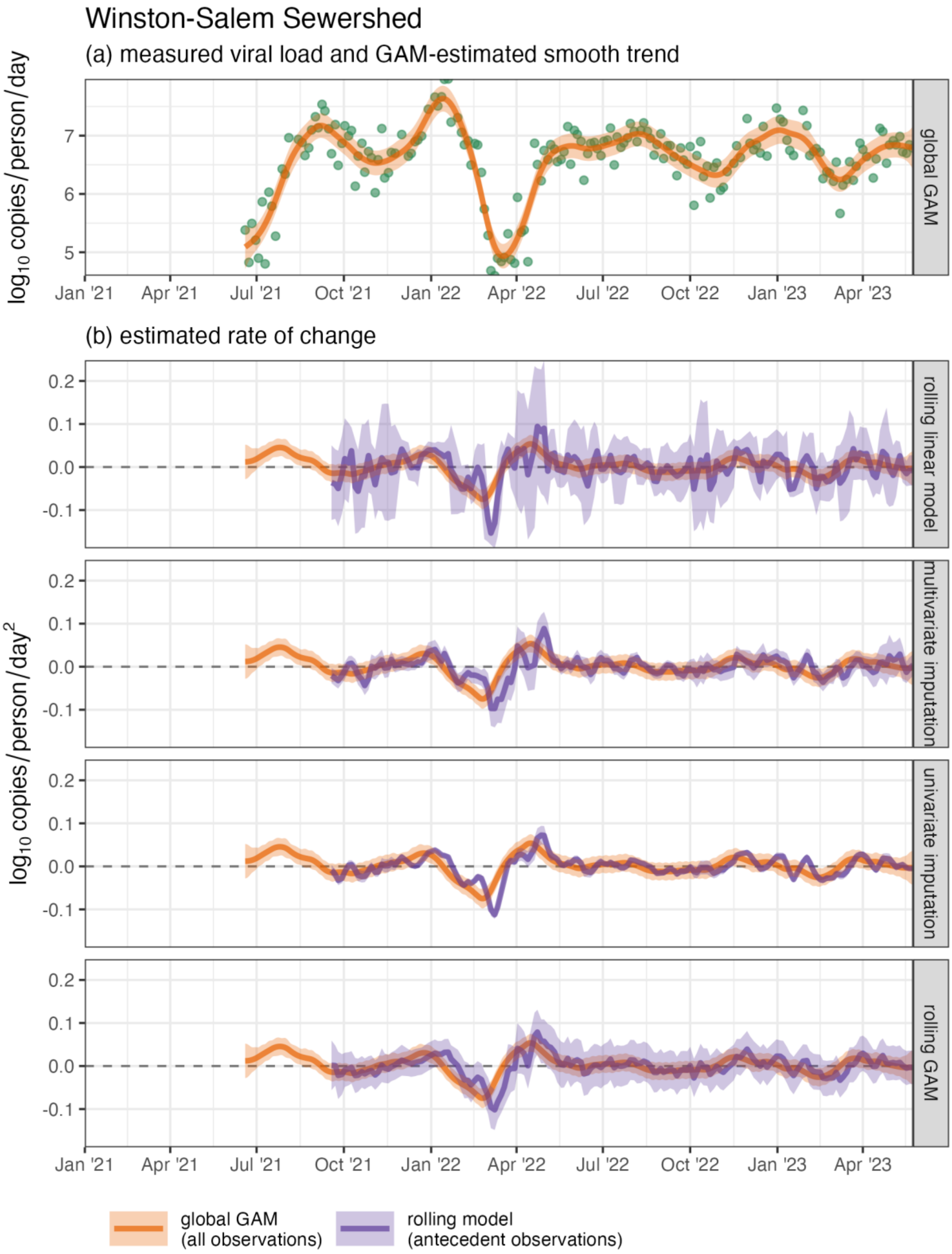

Figure S31. Measured SARS-CoV-2 wastewater per-capita viral load (green points) and estimated mean and 95% CI of the (a) temporal trend and (b) rates of change in the trend for the Winston-Salem sewershed
